# Reliable Contactless Monitoring of Heart Rate, Breathing Rate and Breathing Disturbance During Sleep in Aging: A Digital Health Technology Evaluation Study

**DOI:** 10.1101/2023.10.13.23296936

**Authors:** Kiran K G Ravindran, Ciro della Monica, Giuseppe Atzori, Damion Lambert, Hana Hassanin, Victoria Revell, Derk-Jan Dijk

## Abstract

**Introduction:** Longitudinal monitoring of vital signs provides a method for identifying changes to general health in an individual and particularly so in older adults. The nocturnal sleep period provides a convenient opportunity to assess vital signs. Contactless technologies that can be embedded into the bedroom environment are unintrusive and burdenless and have the potential to enable seamless monitoring of vital signs. To realise this potential, these technologies need to be evaluated against gold standard measures and in relevant populations.

**Methods:** We evaluated the accuracy of heart rate and breathing rate measurements of three contactless technologies (two under-mattress trackers: Withings sleep analyser (WSA) and Emfit QS (Emfit) and a bedside radar: Somnofy) in a sleep laboratory environment and assessed their potential to capture vital signs (heart rate and breathing rate) in a real-world setting. Data were collected in 35 community dwelling older adults aged between 65 and 83 years (mean ± SD: 70.8 ± 4.9; 21 men) during a one-night clinical polysomnography (PSG) in a sleep laboratory, preceded by 7 to 14 days of data collection at-home. Several of the participants had health conditions including type-2 diabetes, hypertension, obesity, and arthritis and ≈49% (n = 17) had moderate to severe sleep apnea while ≈29% (n = 10) had periodic leg movement disorder. The under-mattress trackers provided estimates of both heart rate and breathing rate while the bedside radar provided only breathing rate. The accuracy of the heart rate and breathing rate estimated by the devices was compared to PSG electrocardiogram (ECG) derived heart rate (beats per minute, bpm) and respiratory inductance plethysmography thorax (RIP thorax) derived breathing rate (cycles per minute, cpm). We also evaluated breathing disturbance indices of snoring and the apnea-hypopnea index (AHI) available from the WSA.

**Results:** All three contactless technologies provided acceptable accuracy in estimating heart rate [mean absolute error (MAE) < 2.2 bpm and mean absolute percentage error (MAPE) < 5%] and breathing rate (MAE ≤ 1.6 cpm and MAPE < 12%) at 1 minute resolution. All three contactless technologies were able to capture changes in heart rate and breathing rate across the sleep period. The WSA snoring and breathing disturbance estimates were also accurate compared to PSG estimates (R-squared: WSA Snore: 0.76, p < 0.001; WSA AHI: 0.59, p < 0.001).

**Conclusion:** Contactless technologies offer an unintrusive alternative to conventional wearable technologies for reliable monitoring of heart rate, breathing rate, and sleep apnea in community dwelling older adults at scale. They enable assessment of night-to-night variation in these vital signs, which may allow the identification of acute changes in health, and longitudinal monitoring which may provide insight into health trajectories.

## Introduction

Vital signs measured in clinical practice include heart rate, breathing rate, blood pressure and body temperature. These serve as objective measurements of normal physiological functions and play a fundamental role in assessment of health [1, 2]. With aging, there is an increased incidence of functional limitations and chronic conditions including hypertension, coronary heart disease, stroke, type-2 diabetes, and sleep apnea [3–6]. Standardised, continuous vital signs monitoring systems when implemented for at-home care of older adults, including people living with dementia (PLWD), can serve as an important tool for early identification of changes in health, improve care in older people, and reduce the burden on the healthcare system [7–10]. Commercially available wearable devices (wearables) and contactless technologies (nearables) are increasingly used for home monitoring and have the potential to enable remote health monitoring and promote independent living [11–18]. These technologies offer secure digital infrastructure that allows reliable and seamless transfer of collected data to cloud servers and can facilitate long term remote monitoring opportunities for healthcare.

Wearables are widely used for continuous, community monitoring of heart rate and some have been evaluated in clinical settings although predominantly in younger age groups [17–25]. Although several wearables have been shown to be acceptable for older adults, lower technology adoption rate, user comfort trade off, and burden of maintenance (removal during some daily activities such as showers, periodic recharging, and mobile application use) may make them unsuitable for long term use in PLWD due to their associated behavioural and psychological symptoms [23, 26]. Most wearable devices are limited to activity tracking and heart rate measurement and do not provide other vital signs measures, such as breathing rate, nor can they assess sleep related breathing disorders such as sleep apnea [25, 27].

Contactless technologies can be embedded in the living environment such as under the bed mattress (under-mattress devices or bed sensors) or on the bedside table (e.g., bedside radars) and allow contactless monitoring when the user is in bed. They are powered by the mains and securely stream the collected data wirelessly. They use several different contactless sensing modalities to measure a composite signal (ballistographic signal) containing movements resulting from breathing and cardiac activity to extract vital signs (heart rate and breathing rate) information. The bedside radars use the doppler radar technique while the under-mattress devices employ several different technologies such as electromechanical films and pneumatic sensors [28, 29]. Due to their inconspicuous nature and low maintenance, they do not pose any of the burdens imposed by wearables and are an ideal tool for continuous monitoring of vital signs, behavioural information, and sleep in community dwelling older adult populations, especially in PLWD [13, 14, 30, 31].

To realise the potential of contactless technologies for monitoring vital signs such as heart rate and breathing rate in the community, the accuracy and reliability of their measurements need to be evaluated in relevant populations. While the validity of the heart rate and breathing rate estimates collected from a few contactless technologies has been evaluated in younger populations, to the best of our knowledge, there are no vital signs evaluation studies in older adults (> 65 years) although these devices have been implemented in longitudinal studies [13, 30, 32–34]. Here, we have evaluated the accuracy of heart rate and breathing rate measurements collected from three contactless technologies (a bedside radar and two under-mattress devices) against polysomnography (PSG) electrocardiogram (ECG) derived heart rate and respiratory inductance plethysmography thorax (RIP thorax) derived breathing rate in a laboratory setting. The evaluation addresses aspects of overnight average estimates, ability to capture the overnight trends, variability in heart rate and breathing rate and accuracy at different sleep stages and time resolutions (60, 10 and 1-minute intervals) of estimates. We also discuss the data collection reliability in a home environment and summary estimates of breathing disturbance from the devices. To enhance the relevance of this study we applied liberal inclusion/exclusion criteria for the participant selection such that several participants had co-morbidities which is representative of the general older population.

## Methods

### Cohort characteristics

The study data were collected at-home for a period of 7 to 14 days followed by an overnight laboratory session (with full polysomnography (PSG)) at the Surrey Sleep Research Centre in two cohorts (Cohort 1 [Jan. – Mar. 2020] and Cohort 2 [Jun. – Nov. 2021] including 18 and 17 participants, respectively). The participant group consisted of 35 individuals (21 men) between the ages of 65 and 83 (70.8 ± 4.9 [mean ± SD]). The participants were identified and recruited through the Surrey Clinical Research Facility (CRF). To ensure the ecological validity of the collected data in this population, participants with stable co-morbidities such as hypertension, type-2 diabetes, arthritis etc were included in the study provided that their co-morbidity and concomitant medications were stable and won’t pose safety risks if they participate. Eligible participants had to be able to independently perform activities of daily life and comply with study procedures. Participants were provided with detailed information about the study and provided written informed consent before any study procedures were performed. The study received a favourable opinion from the University of Surrey Ethics committee (UEC-2019-065-FHMS) and was conducted in line with the Declaration of Helsinki and the principles of Good Clinical Practice. A detailed description of the inclusion exclusion criteria can be found in our previous publications in which we evaluate the ability of these technologies to estimate sleep timing in the home environment and sleep stages as defined by polysomnography [35, 36].

### Study protocol

The under-mattress devices (Withings sleep analyser [WSA, Withings, France] and Emfit QS [Emfit Ltd, Finland]) were deployed both in the laboratory and at-home while the bedside radar (Somnofy, VitalThings, Norway) was only used in the laboratory. The participants also used an actigraphy device (Actiwatch spectrum [AWS], Philips Respironics) and maintained a consensus sleep diary at-home [37]. During the home deployment period, the contactless technologies did not require any manual intervention or maintenance by the participants and transmitted the data automatically via Wi-Fi. To ensure anonymity of the data, no personally identifiable participant information was added to the device applications and a portable Wi-Fi router was used for data transfer. After the home data collection period, one overnight full clinical PSG recording was conducted that included an extended time in bed of 10 hours. The WSA was used both in cohort 1 and 2 while the Somnofy and Emfit were deployed only in cohort 2. The data from the contactless technologies were collected simultaneously along with PSG in laboratory and with AWS and sleep diary at-home. Empatica E4 (Empatica Srl, Milan, Italy), a wrist worn device that collects activity and photoplethysmography was also deployed during the laboratory session and is used as an activity reference in this work [26, 38].

### The reference vital signs data

During the in-laboratory session, PSG was collected using the SomnoHD system (SOMNOmedics GmbHTM, Germany). The collected data included electroencephalography (EEG; 256 Hz; F3-M2, C3-M2, O1-M2, F4-M1, C4-M1 and O2-M1), ECG (modified lead II sub clavicular electrode placement; 256 Hz), RIP thorax and abdomen (128 Hz), photoplethysmography (PPG, 128Hz), electromyography (EMG; 256Hz, both submental and limb) and electrooculography (EOG; 256Hz; E2-M1 and E1-M2). Additionally, snore sensor (256 Hz) and air flow via nasal cannula and flow thermistor (128 Hz) data were also collected. Sleep was scored at 30 s intervals in the DOMINO software environment as per American Academy of Sleep Medicine (AASM) guidelines by two independent scorers (a Registered Polysomnographic Technologist™ [RPSGT] and a trained scorer) and a consensus hypnogram was generated [38]. The sleep hypnogram contains five stages: rapid eye movement (REM), stage N1 of non-REM sleep (N1), stage N2 of non-REM sleep (N2), and stage N3 of non-REM sleep (N3), and wake. The apnea-hypopnea index (AHI; number of apnea/hypopnea per hour) and period limb movement index (PLMI; number of period limb movement events per hour) were determined by the RPSGT using scoring rules recommended by AASM. An apnea was scored when there was a ≥ 90 % drop in airflow lasting for at least 10 s while a hypopnea was scored using the 3 % drop in oxygen saturation and/or an arousal in the electroencephalogram criteria. Severity of apnea was determined using the following thresholds as per AASM guidelines: AHI-< 5 normal, 5 to < 15 mild apnea, 15 to <30 moderate apnea and ≥30 considered severe. A periodic limb movement event was scored when at least four consecutive limb movements occurred, each separated from the preceding limb movement by at least 5 s, but not more than 90 s apart. PLMI of more than 15 is used as the cut-off for the presence of periodic limb movement disorder [39, 40]. Additionally, participants with cardiac arrythmia were identified using the arrythmia index generated by the DOMINO software and verified by visual inspection of the record.

The PSG data were exported as standard edf+ files along with recording markers and the consensus hypnogram. The ECG from the PSG was used for the extraction of the heart rate reference data while RIP thorax was used as the breathing rate reference data. For one of the participants where RIP thorax was unavailable, RIP abdomen was used to create the breathing rate reference data. MATLAB® 2021b was used for all the data analysis reported. The RR intervals used for the computation of the heart rate were derived from the ECG using the PhysioNet Cardiovascular Signal Toolbox and a well evaluated beat detection toolbox [41, 42]. This beat-to-beat information was used to estimate reference heart rate (beats per minute, bpm) at 30s intervals which is the same as the PSG hypnogram. For extracting the breathing rate (cycles per minute, cpm) from the RIP thorax signal at 30 s intervals, the RRest package was used [43, 44].

### Contactless technologies: Data overview

The WSA and Somnofy data (json format) were downloaded using the respective manufacturer’s application programming interface (API) while the Emfit data (csv format) were downloaded from the manufacturer’s web interface. All the compared contactless technologies (WSA, Emfit and Somnofy) provided breathing rate data while only the under-mattress devices (WSA and Emfit) provided heart rate data.

The devices provided vital signs (heart rate and breathing rate) data and sleep hypnograms at different resolutions (WSA: 60 s; Emfit and Somnofy: 30 s). Throughout this article, we have used the term vital signs to denote heart rate and breathing rate and vice versa. The WSA and Somnofy heart rate and breathing rate estimations were available at the 60 s and 30 s resolutions (same as the respective device hypnogram resolution) while the Emfit estimated heart rate at 4 s intervals. These 4 s estimates were averaged to generate estimates at 30 s intervals to match the Emfit sleep label intervals. To allow data analysis relative to local time, daylight savings correction was applied to the UTC timeseries generated by the devices. The sleep hypnograms generated by the devices contain four stages: deep sleep (DS = N3), light sleep (LS = N2/N1), REM and wake.

Apart from heart rate and breathing rate, Emfit generates continuous heart rate variability and activity measures while Somnofy provides estimates of movement and environmental variables such as ambient light, sound, temperature, pressure, humidity, and indoor air quality which were out of scope for this evaluation and are not discussed here.

Both at home and in the laboratory, all devices were connected to the same network and the devices used manufacturer’s time synchronisation protocol such as network time protocol (NTP) to timestamp the data. Although this ensured that the devices were synchronised to local time, we performed another synchronisation step to allow accurate comparison of the data between the devices. The device vital signs measures were aligned to the PSG reference vital signs estimates via cross correlation between the device and PSG vital signs and hypnograms and the lag (within a 5 min window) that provided the best alignment of the both the vital signs data and hypnograms was then applied. The WSA data were converted from 60 s to 30 s intervals by up sampling. Epochs in the PSG and device hypnograms that were scored as artefacts/no presence were excluded from the assessment.

### Vital signs assessment

The evaluation of the epoch-by-epoch (EBE) heart rate and breathing rate data collected by the contactless technologies was performed against reference estimates derived from PSG ECG and RIP thorax. The accuracy and reliability of the heart rate and breathing rate estimates were performed at different levels of time resolution to determine use cases in which the contactless technologies can be employed. These include accuracy assessment of overnight average estimates, ability to capture overnight trends, variability in vital signs, and accuracy in different sleep stages and at different time resolutions (60,10, and 1 minute intervals) of estimates. All the laboratory data analyses were performed over the total recording period of the PSG. At all temporal resolutions of comparisons only complete/valid pairs of estimates were used.

#### Performance measures

For assessing the accuracy of the vital signs estimates (heart rate and breathing rate), mean absolute error (MAE) and mean absolute percentage error (MAPE) were used as the primary metrics. MAE and MAPE are used to measure the error in the estimate between the device and the PSG reference vital signs. The Bland-Altman metrics such as minimum detectable change (MDC), bias and limits of agreement (LoA) were also computed to provide an overview of the agreement of the estimates and to allow comparison with evaluations reported in the literature [45, 46]. All measures are reported with 95 % confidence intervals. MDC is the smallest change in the estimate that can be detected by the device that exceeds the measurement error. We have used intraclass correlation with two-way random effects (ICC) to measure the reliability of measurement and standardised absolute difference (SAD), a directionless Cohen’s D described in [36, 47], for measuring the dispersion in the bias. Apart from the above metrics, coefficient of determination (r^2^, measure of how close the measured estimates are to the reference computed using simple linear regression) was also used for the concordance analysis. For estimating the significance of differences between vital signs during different sleep stages, devices, and time course we used ANOVA followed by linear mixed effects models with the different groups (devices, sleep stages and time) as fixed effects (with interactions) and participant as random effect.

#### Acceptable agreement for heart rate

The satisfactory level of agreement between the PSG reference heart rate and the device determined heart rate was set to an error of 10 % or ± 5 bpm as recommended by the Association for the Advancement of Medical Instrumentation (AAMI, ANSI/AAMI 2002) [42, 48].

#### Acceptable agreement for breathing rate

For breathing rate, to the best of our knowledge, there is no device specific satisfactory level of agreement discussed in literature. Hence, the agreement in breathing rate estimates between human observers is used to set the acceptable level for our evaluation. We set the permissible level of error in breathing rate estimation to be ± 4 cpm as reported by Lim et al [49].

### Breathing disturbance estimates

Apart from EBE heart rate and breathing rate, WSA also provides a ‘Snoring’ signal which is a binary variable depicting snore presence detected by the device. In addition, the WSA also provides summary estimates of snoring duration (WSA snore), breathing disturbance intensity (WSA BDI) and AHI (WSA AHI). The Emfit and Somnofy devices do not generate breathing disturbance measures. Only complete/valid pairs of estimates were used in analysing the summary measures. For WSA BDI and AHI estimates, we explored the relation between them and the concordance of these WSA estimates to the PSG AHI in the laboratory.

For the WSA snore estimates, we explored the concordance of the all-night snore duration estimated by the PSG snore sensor (i.e., PSG snore microphone placed on the side of the neck) and the WSA. The PSG snore sensor data were scored by the Somnomedics DOMINO software using 30dB as snore amplitude threshold and a minimum snore duration of 300 ms. We further explored the differences in the snore intensity as measured by the PSG snore sensor for the epochs determined to contain snore events by the WSA followed by an exploration of distribution of the snore events during the different sleep stages.

## Results

### Characteristics of the study population

More than half of the study participants (n = 20; > 57%) reported co-morbidities including type-2 diabetes, obesity, arthritis, and hypertension, with concomitant medication. In this study population, the mean heart rate (mean ± standard deviation) was 62.2 ± 8.9 bpm (Men [n = 21]: 60.6 ± 9.3 bpm; Women [n = 14]: 64.4 ± 7.8 bpm) and mean breathing rate was 14.7 ± 2.9 cpm (Men: 14.6 ± 2.9 cpm; Women: 14.7 ± 3.0 cpm), as assessed from the overnight laboratory polysomnography. The average body mass index (BMI) of the participants was 27.0 ± 4.8 kg/m^2^ with 17 % (n = 6, BMI > 30) being obese. The mean systolic and diastolic blood pressures measured during screening were 148.7 ± 16.2 mmHg and 87.0 ± 9.6 mmHg, respectively. During the clinical PSG it emerged that 95 % (n = 33) of the participants in the study had some degree of apnea. Of the participants with apnea, eight had severe (≈23%, apnea hypopnea index (AHI): > 30), nine had moderate (≈26 %, AHI: 15 to < 30) and 16 had mild apnea (≈46%, AHI: 5 to < 15). Ten (28.57%) participants had PLMI > 15 which is similar to the prevalence of periodic limb movement syndrome in community dwelling elderly [50, 51]. Some form of cardiac arrhythmia was found in ≈ 51 % (n = 18) of the participants with ≈ 29% (n = 10) of them also having severe / moderate apnea. A detailed description of the population characteristics can be found in [35].

### Overview of vital signs data

#### Example case

An example 15 day (14 days at-home and one day in laboratory) vital signs data collected by WSA is shown in Figure 1. The participant had moderate apnea with an AHI of 24.1 events/hour as determined by the clinical PSG during the laboratory visit (day 0). The raster plot (Figure 1 a) shows the heart rate, sleep/wake stage, and breathing rate as estimated by the WSA. The vital signs data were available when the participant was in bed at night and during daytime in bed periods which were also reported as naps by the participant. This participant had an irregular nocturnal bed timing with an average time in bed during the nocturnal period of 10 hours 8 minutes at home. Mean nocturnal heart rate and breathing rate varied across nights. The WSA AHI showed night –to-night variability (see Figure 1 b) and the WSA AHI during the laboratory visit was 25 events/hour which was close to the PSG AHI value. The heart rate showed a trend across the nocturnal sleep period with higher heart rate at the beginning of the nocturnal sleep period and a lower heart rate just before the end of the sleep period.

**Figure 1.**
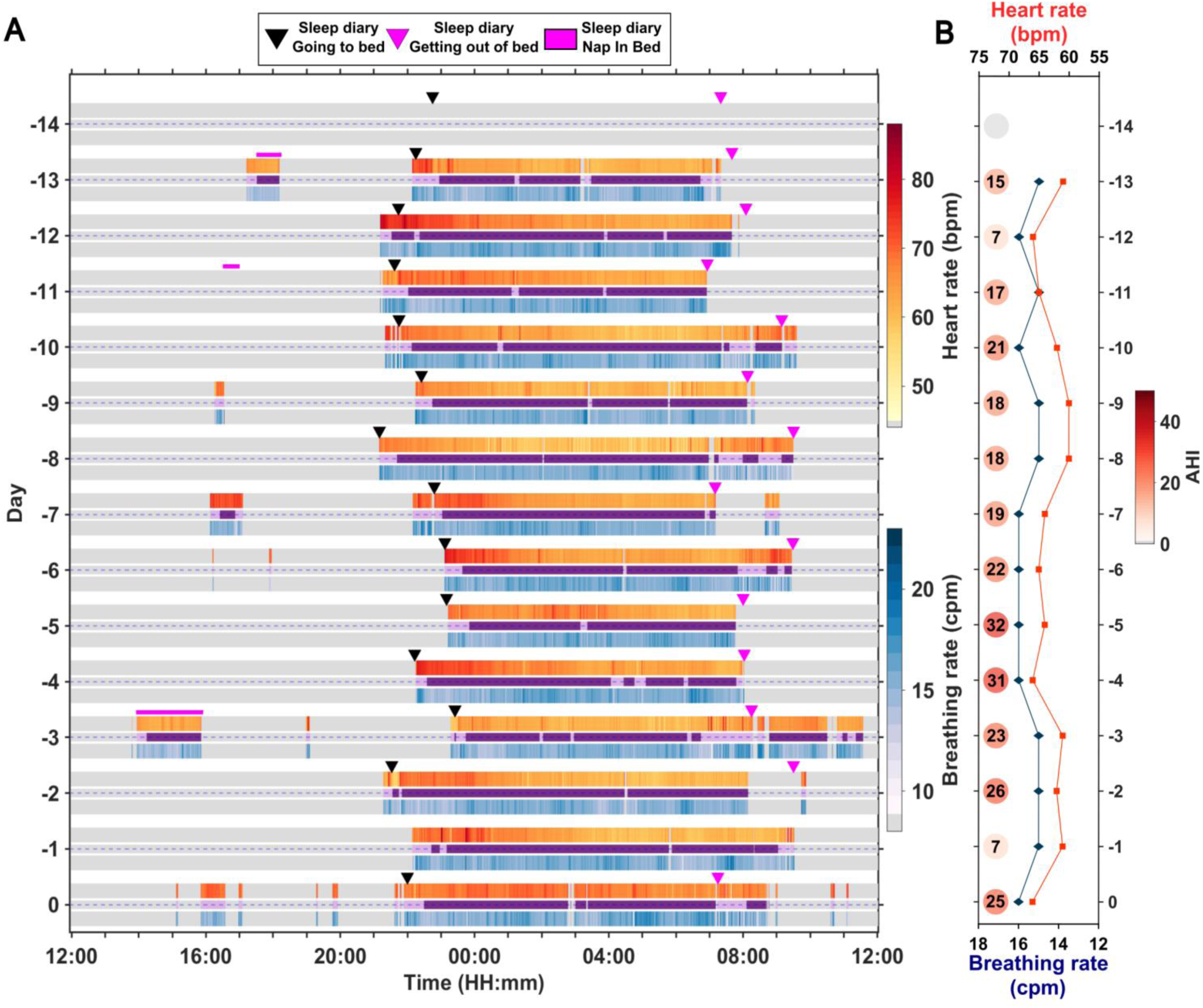
Vital signs collected at home and in the laboratory from a male participant between the ages of 65 and 70. **A**. Raster plot showing the heart rate (beats per minute, bpm) and breathing rate (cycles per minute, bpm) as detected by the Withings Sleep Analyser (WSA) along with device detected sleep (or) wake period and sleep diary information. The vital signs data were available only when the participant was in bed. The days –14 to –1 depict the data collected at home while day 0 depicts data collected in the sleep laboratory. The darker, purple-coloured regions denote sleep while the lighter regions denote wake as identified by WSA. The grey areas represent out of bed periods, i.e., periods during which the device did not record data. **B.** Estimates of mean heart rate, mean breathing rate and apnea-hypopnea index (AHI, depicted as circles adjacent to the time courses) during the night as determined by the device. The first day of data from the WSA at-home was lost.

Figure 2 shows the contactless technology vital signs data collected alongside PSG reference data during the laboratory visit for the participant depicted in Figure 1. The PSG heart rate and breathing rate both show changes as the hypnogram transitions between different stages of sleep with higher variability during wake and rapid eye movement (REM) sleep and lower variability during non-rapid eye movement (NREM) sleep. The differences in the data resolution (WSA: 60s; Emfit and Somnofy: 30s) between the contactless technologies can be seen from the plots. The WSA vital signs estimates provided by the device are rounded to the nearest integers leading to a more discretised vital signs data. The trends in the heart rate and breathing rate data recorded by the PSG and the contactless technologies are more similar during the sleep periods compared to the wake periods. The WSA had the most similarity to the PSG followed by Somnofy and Emfit. The Emfit heart rate had large deviations compared to the PSG estimates during wake periods. The Somnofy on the other hand had a number of missing estimates of breathing rate during many of the wake epochs determined by the device and these missing vital signs epochs also coincided with periods of higher activity as detected by a wrist worn activity device (Empatica E4). Snoring as detected by the WSA followed a pattern which closely followed the snoring signal detected by PSG with some disagreement in the snore event detection (see Figure 2).

**Figure 2.**
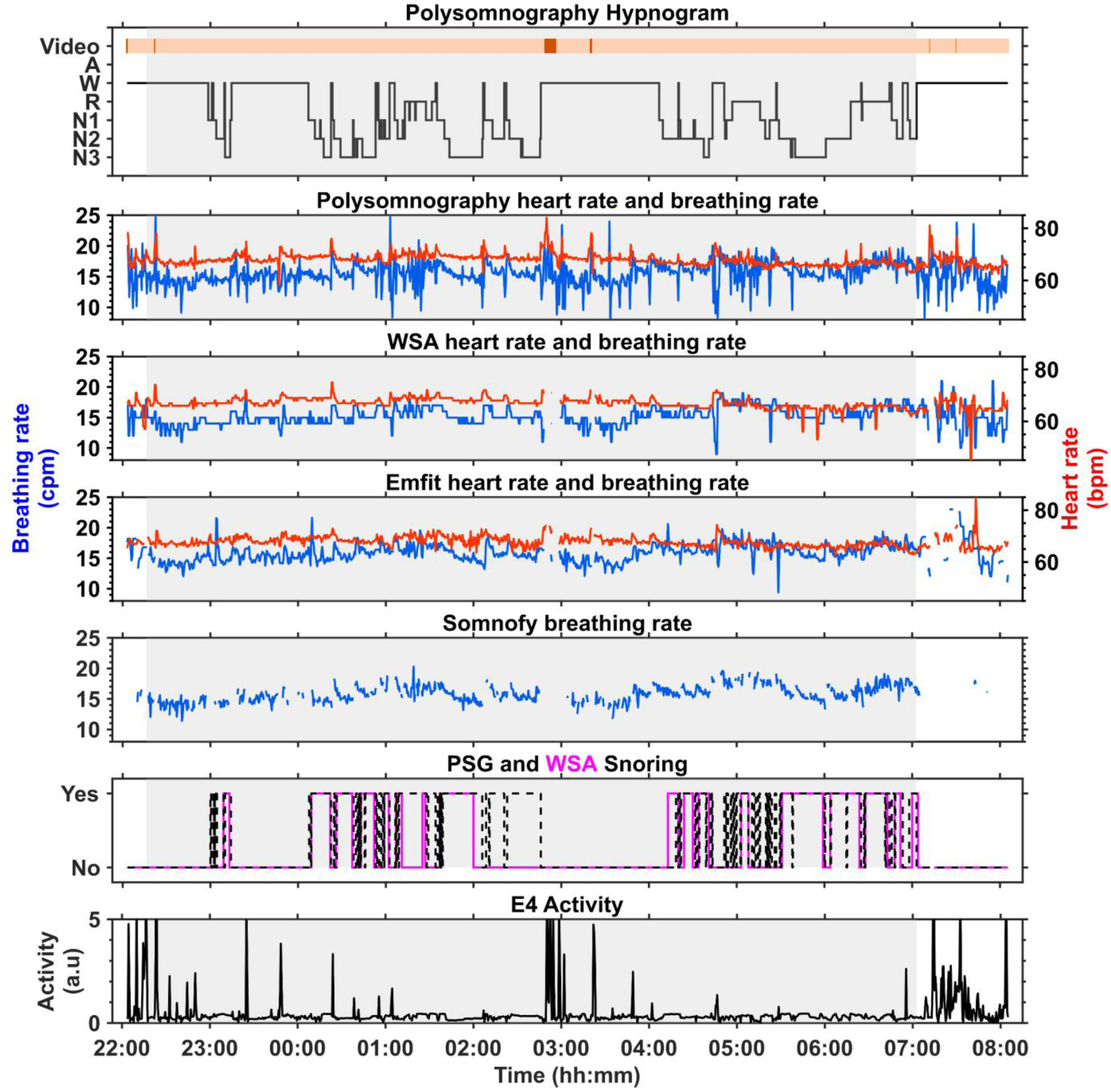
Vitals signs data collected from three contactless technologies simultaneously alongside polysomnography (PSG) in the laboratory for the participant depicted in Figure 1. The PSG consensus hypnogram is depicted at the top followed by the heart rate ([**Red**] beats per minute, bpm) and breathing rate ([**Blue**] cycles per minute, cpm) from PSG and contactless devices. The activity data is from Empatica E4 device. The grey regions in the plots correspond to the lights off period.

#### Summary of collected data

In the laboratory study, all 35 PSGs (ground truth/reference data, cohort 1: n = 18 and cohort 2: n = 17) were available. The total number of nights of data collected in the laboratory for each of the three contactless technologies were WSA = 35, Emfit = 16 and Somnofy = 17. One night of data was lost due to device malfunction for Emfit. At-home, A total of 401 days of data were collected across the 35 participants with 321 days of data available for WSA (cohort 1: n = 10; cohort 2: n = 17) and 228 days of data available for Emfit (cohort 2: n = 17). At-home, for WSA, portions of data from eight participants were lost in cohort 1 due to deployment errors and Wi-Fi dropouts with a data loss of 3.3 % (11 days out of 332). For Emfit the data loss was 4.2 % (10 days lost out of 238).

In the laboratory, the range of the heart rate estimated ([min, max]) by the contactless technologies were, WSA: [40, 90] and Emfit: [40, 135], bpm whereas for breathing rate the ranges were WSA: [8, 35], Emfit: [6, 30] and Somnofy: [6, 30] cpm. The WSA breathing rate range was also smaller than the ranges of Emfit and Somnofy. The under-mattress device generated vital signs (both heart rate and breathing rate) data for 100 % of the in-bed periods. Somnofy bedside radar, on the other hand generated breathing rate data less continuously resulting in a data unavailability of 32.21 % of the in-bed period (in-laboratory). Most of these missed breathing rate epochs were found to be in the Somnofy predicted wake state (% total breathing rate epochs unavailable per label: Wake = 62.39 %; REM = 12.73 %; LS = 23.10 %; DS = 1.78 %).

### All night vital signs

Concordance between the nightly average heart rate and breathing rate estimates of the contactless technologies against PSG are shown in Figure 3 and Table 1. The WSA (MAPE: 3.28 %; ICC: 0.87) had a lower level of agreement compared to Emfit (MAPE: 1.83 %; ICC: 0.96) and this was due to an outlier participant with severe arrythmia (see the outlier in Figure 3).When the outlier was removed WSA (WSA*, MAPE: 1.87 %; ICC:1.0) had an agreement with the PSG which was similar to that of Emfit. The MDC was higher for Emfit (3.25 bpm) compared to WSA* (1.17 bpm) which can be seen from the higher dispersion in the Emfit estimates (Figure 2 a). For the breathing rate, Somnofy (MAPE: 4.64 %; ICC: 0.82) had a high agreement followed by WSA (MAPE: 6.29 %; ICC: 0.78) and Emfit (MAPE: 5.46 %; ICC: 0.76). The MDC follows the agreement results, with Somnofy (1.98 cpm) having a somewhat lower value compared to WSA (2.08 cpm) and Emfit (2.21 cpm).

**Figure 3.**
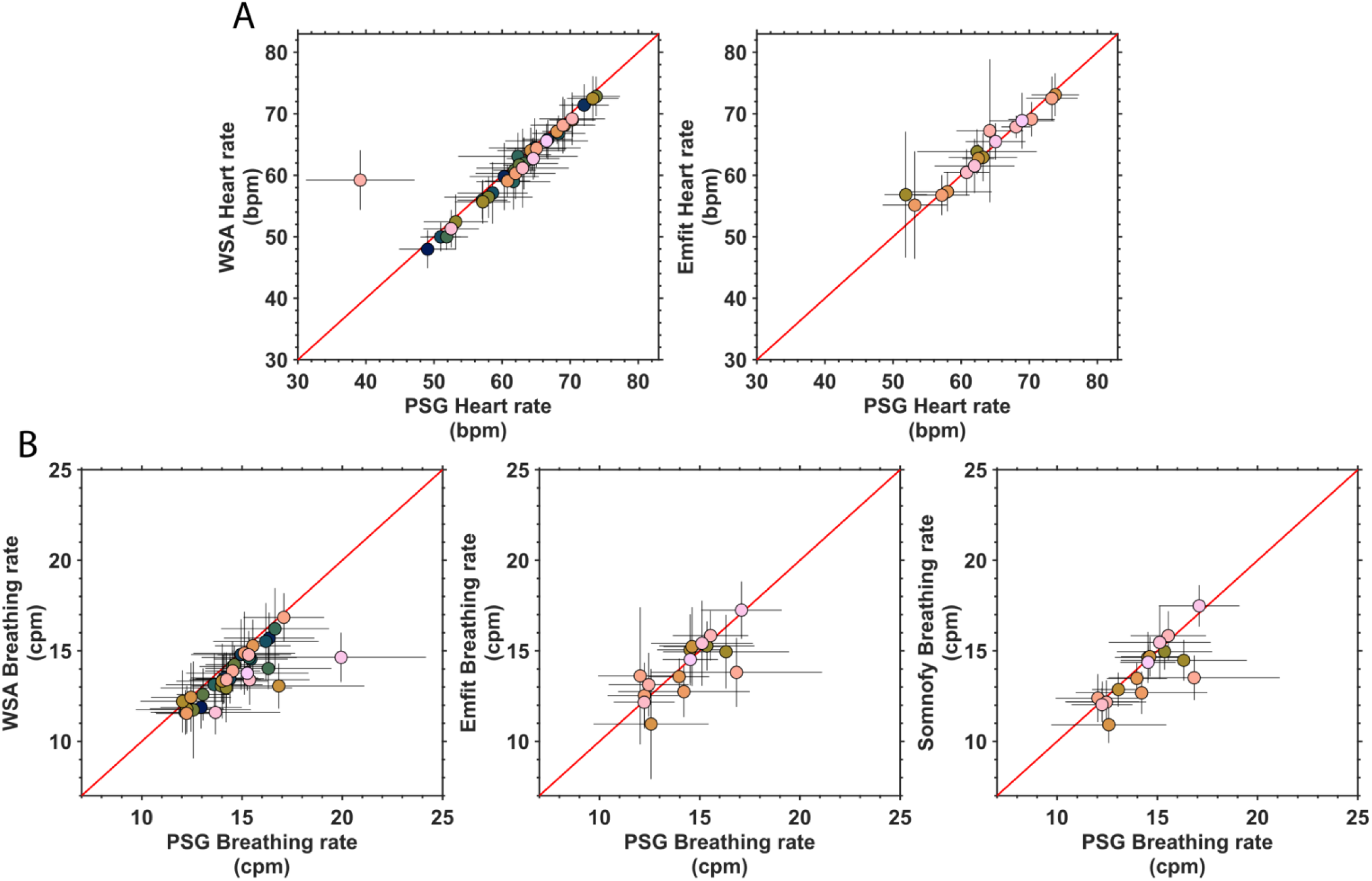
Association between vital signs estimated by three contactless devices and estimates from polysomnography (PSG) averaged across the night while sleeping in the sleep laboratory. **A**. Heart rate (beats per minute, bpm) and **B.** Breathing rate (cycles per min, cpm). The error bars represent the standard deviation of the estimate within participants. The PSG heart rate and breathing rate are derived from electrocardiogram (ECG) and respiratory inductance plethysmography thorax (RIP thorax) respectively. The number of participants (=nights) available for the devices are WSA [n=34], Emfit [n=16] and Somnofy [n=17].

**Table 1.** All night average of vital signs and their agreement metrics.

| Vital Signs |  | Device<br>Mean<br>(SD) | PSG<br>Mean<br>(SD) | Bias<br>[95% CI] | LoA Lower<br>bound<br>[95% CI] | LoA<br>Upper<br>bound<br>[95% CI] | MD<br>C | MAE<br>[95%<br>CI] | SAD<br>[95%<br>CI] | MAPE<br>[95%<br>CI] | ICC<br>[95%<br>CI] |
| --- | --- | --- | --- | --- | --- | --- | --- | --- | --- | --- | --- |
| Heart rate | WSA | 61.66<br>(6.51) | 62.16<br>(7.52) | -0.5 (3.63)<br>[-1.75,<br>0.74] | -7.61<br>[-9.76, -<br>5.46] | 6.61<br>[4.46,<br>8.75] | 7.1<br>1 | 1.69<br>[2.8,<br>0.58] | 0.24<br>[-0.09,<br>0.58] | 3.28<br>[0.40,<br>6.16] | 0.87<br>[0.75,<br>0.93] |
|  | WSA* | 61.73<br>(6.6) | 62.84<br>(6.46) | -1.11 (0.6)<br>[-1.31, -0.9] | -2.28<br>[-2.64, -<br>1.92] | 0.07<br>[-0.29,<br>0.43] | 1.1<br>7 | 1.15<br>[1.33,<br>0.97] | 0.18<br>[-0.16,<br>0.52] | 1.87<br>[1.56,<br>2.17] | 1<br>[0.99,<br>1] |
|  | Emfit | 63.85<br>(5.66) | 63.42<br>(6.47) | 0.44 (1.66)<br>[-0.45,<br>1.32] | -2.81<br>[-4.35, -<br>1.27] | 3.68<br>[2.14,<br>5.22] | 3.2<br>5 | 1.08<br>[1.78,0<br>.39] | 0.18<br>[-0.33,<br>0.7] | 1.83<br>[0.52,<br>3.13] | 0.96<br>[0.9,<br>0.99] |
| Breathing<br>Rate | WSA | 13.66<br>(1.38) | 14.61<br>(1.69) | -0.95 (1.06)<br>[-1.32, -<br>0.59] | -3.04<br>[-3.66, -<br>2.41] | 1.13<br>[0.5, 1.75] | 2.0<br>8 | 0.97<br>[1.33,<br>0.6] | 0.63<br>[0.3,<br>0.97] | 6.29<br>[4.31,<br>8.27] | 0.76<br>[0.58,<br>0.87] |
|  | Emfit | 14.13<br>(1.6) | 14.35<br>(1.67) | -0.22 (1.13)<br>[-0.82,<br>0.38] | -2.43<br>[-3.48, -<br>1.38] | 1.99<br>[0.94,<br>3.03] | 2.2<br>1 | 0.78<br>[1.22,0<br>.34] | 0.49<br>[-0.02,<br>1.01] | 5.46<br>[2.55,<br>8.36] | 0.76<br>[0.44,<br>0.91] |
|  | Somnofy | 13.77<br>(1.68) | 14.28<br>(1.65) | -0.51 (1.01)<br>[-1.02,<br>0.01] | -2.48<br>[-3.38, -<br>1.58] | 1.47<br>[0.57,<br>2.37] | 1.9<br>8 | 0.69<br>[1.14,<br>0.24] | 0.43<br>[-0.07,<br>0.93] | 4.64<br>[1.79,<br>7.5] | 0.82<br>[0.56,<br>0.93] |

Note. Metrics of agreement for overall heart rate of the devices against the electrocardiogram (ECG) (included in the polysomnography, PSG) estimates (beats per minute, bpm). The values shown are the mean followed by the standard deviation (values reported in parentheses) and 95% confidence interval (values reported in squared brackets). WSA\* depicts the outlier removed WSA data. The metrics include: Bias – difference in measurement between the Device and PSG-ECG (Device vs PSG-ECG); Lower and Upper bounds of the Bias; Minimum detectable change (MDC) – smallest detectable change independent of measurement error (half of Bland Altman agreement width); Standardized absolute difference (SAD) – directionless version of Cohen's d; Mean absolute percentage error (MAPE) – mean error in measurement; Intraclass correlation with two-way random effects (ICC) – measure of measurement reliability. The number of participants contributing to each of the devices are WSA [n=35], WSA\* [n=34], Emfit [n=16] and Somnofy [n=17].

### Vitals signs during different vigilance states

We investigated the agreement of the vital signs estimated by the contactless technologies to the PSG reference during the different vigilance states of the consensus PSG hypnogram. The distribution of the vital signs for the different vigilance states is provided in Supplemental Figure 1. The heart rate and breathing rate estimates of the both PSG and contactless devices were not normally distributed (via Kolmogorov-Smirnov test). The difference between the mean heart rate estimated by the devices and PSG was less than 2.5 bpm across all sleep stages. The difference between the mean breathing rate estimated by the devices and PSG was less than 1.5 cpm across all sleep stages. Overall, the concordance between the estimates of both heart and breathing rate provided by the contactless technologies and PSG was good across all vigilance states (See supplemental Figure 2 and supplemental Table 1 and 2).

### Time course of vital signs during sleep

Figure 4a shows the time course of the heart rate and breathing rate estimated by the contactless devices and PSG over the PSG lights off period in-laboratory (Figure 4 a) Fig 4b shows these time course for the sleep diary defined lights-off periods recorded at-home (Figure 4 b). The heart and breathing rate average were computed over the epochs during which participants were asleep as detected by the PSG hypnogram for the laboratory data and as detected by the sleep scoring algorithms of the devices for the at-home data.

**Figure 4.**
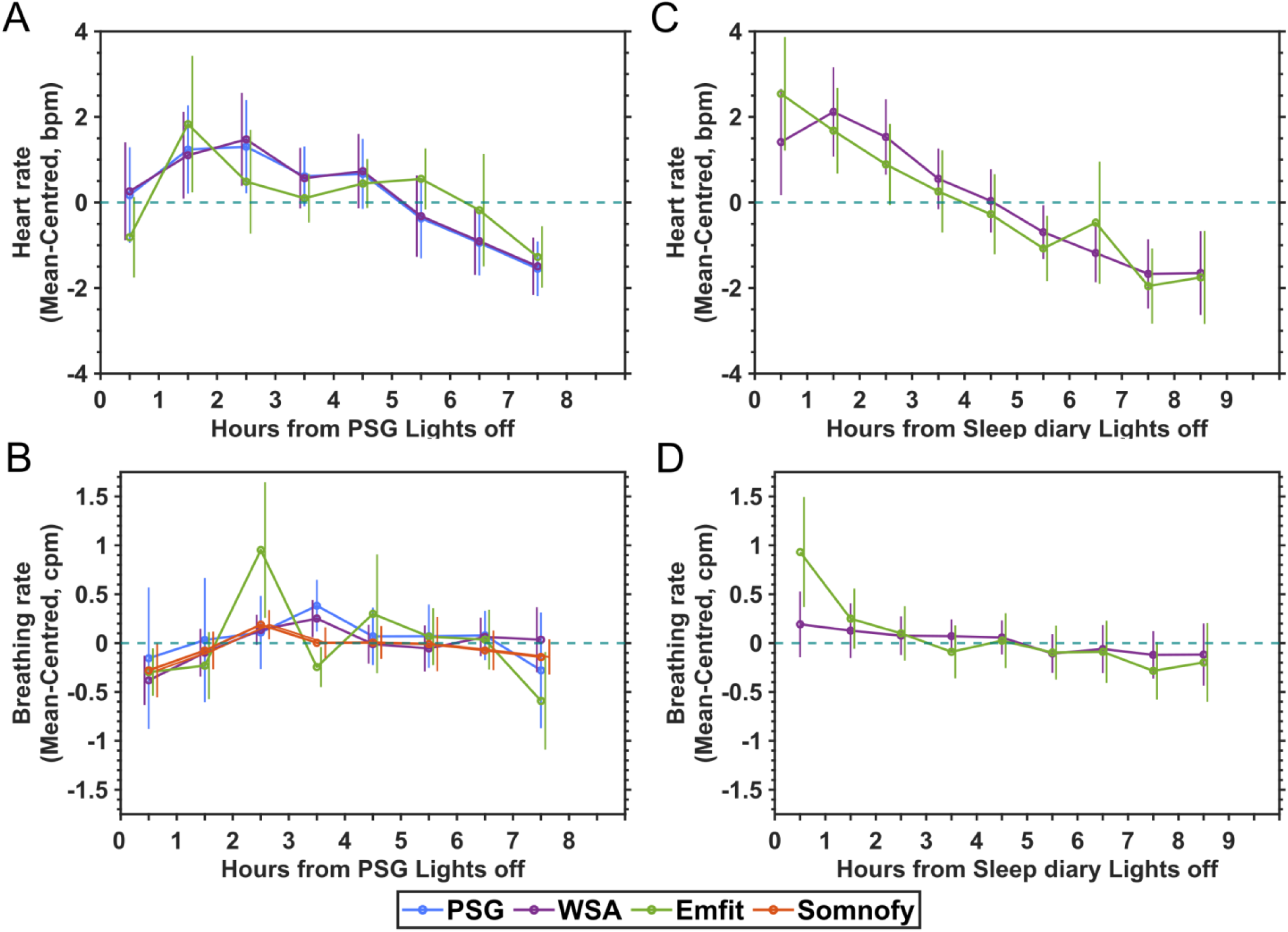
Time course of vital signs during sleep. and B. In-laboratory **C and D.** at home. The vital signs data were mean centred and averaged during sleep and plotted per hour starting from the onset of the lights off period. The error bars represent the standard deviation of the estimate and are shifted along the x axis to improve visibility. For all devices, the consensus PSG hypnogram was used to select vital signs data during sleep in the laboratory and the hypnograms as generated by the devices were used for the analyses of the data collected at home. The number of participants available for each of the devices: In-laboratory – PSG [n=35, 35 nights], WSA [n=34, 34 nights], Emfit [n=16, 16 nights] and Somnofy [n=17, 17 nights]; At home – WSA [n=27, 295 nights] and Emfit [n=17, 213 nights].

Both in the laboratory and at-home, the heart rate started close to or above the mean, gradually decreases over the night and reached the nightly minimum in the second part of the sleep period before increasing again. The similarity between the contactless technology and PSG heart rate hourly timeseries in-laboratory was determined using MAPE. The WSA (MAPE: 14.42 %) closely follows the PSG while the Emfit (MAPE: 144.34 %) is less similar.

The breathing rate trends at-home were similar to that of the heart rate starting above the overnight mean gradually decreasing to a lower value closer to the end of the night. In the laboratory, the breathing rate hourly estimates were fluctuating and did not show any clear trend. Both WSA (MAPE: 129.95 %) and Somnofy (MAPE: 130.01 %) had the highest similarity with PSG while Emfit had the lowest similarity (MAPE: 295.76 %).

### Effect of temporal resolution on the vital signs accuracy

To examine the effect of the length of the time period over which the vital signs are computed, we averaged the heart rate and breathing rate estimates over 60 minutes, 10 minutes and 1 minute and computed the agreement with the corresponding PSG reference estimates. We examined the cumulative distribution function (CDF) of the MAE at these resolutions to better characterise the estimation error. The CDFs are depicted in Figure 5 while more detailed scatter plots and associated agreement measures are provided in Supplemental Figure 3 and Table 2 and 3.

**Figure 5.**
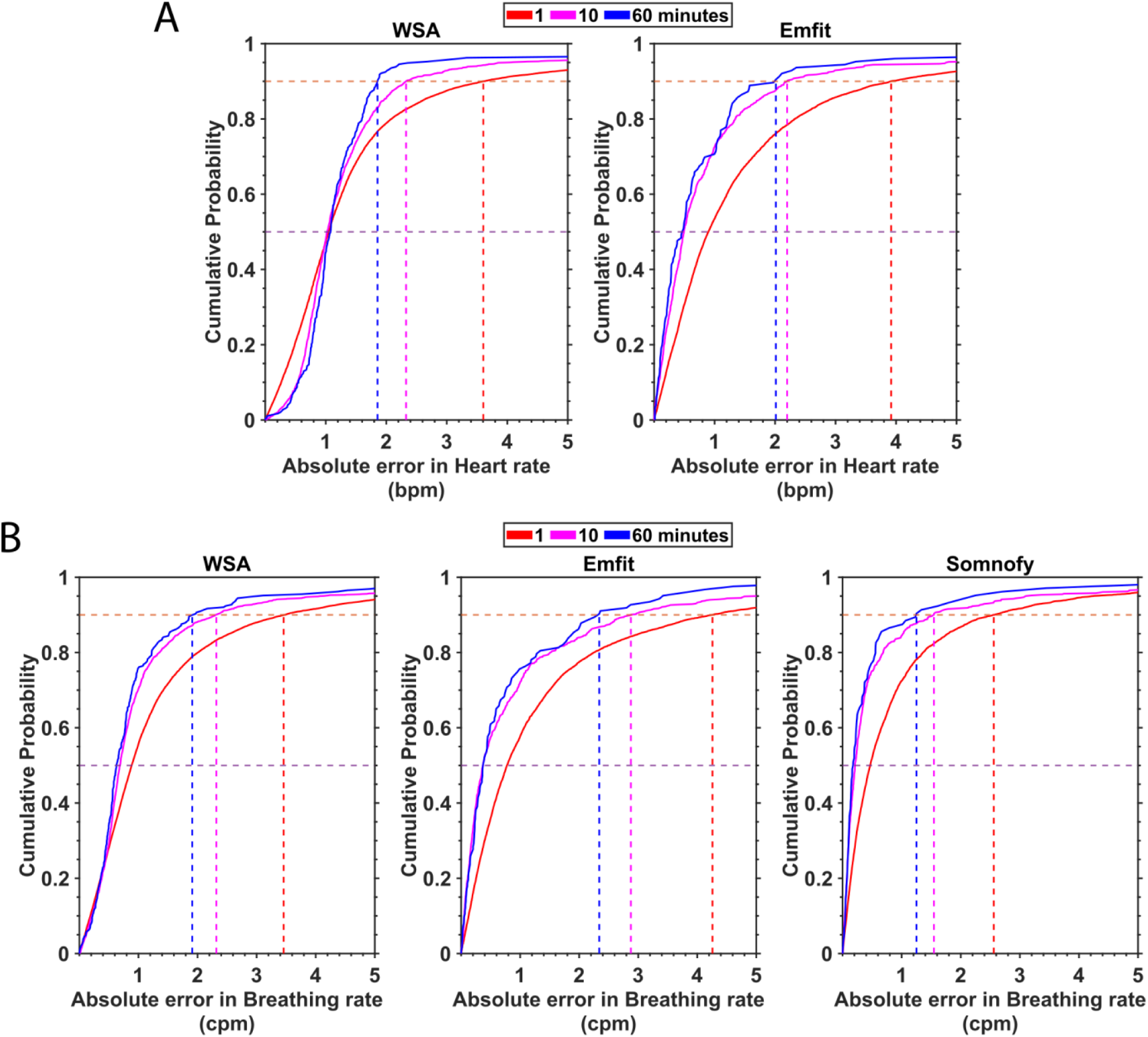
Effect of time window over which vital signs are estimated on device measurement error. **A**. Heart rate (beats per minute, bpm) and **B.** Breathing rate (cycles per minute, cpm). The cumulative density function of the absolute error is represented for each of the devices for the window lengths 1 min,10 min and 60 min. The median (50^th^ percentile) and the 90^th^ percentile are represented by horizontal lines.

**Table 2.**
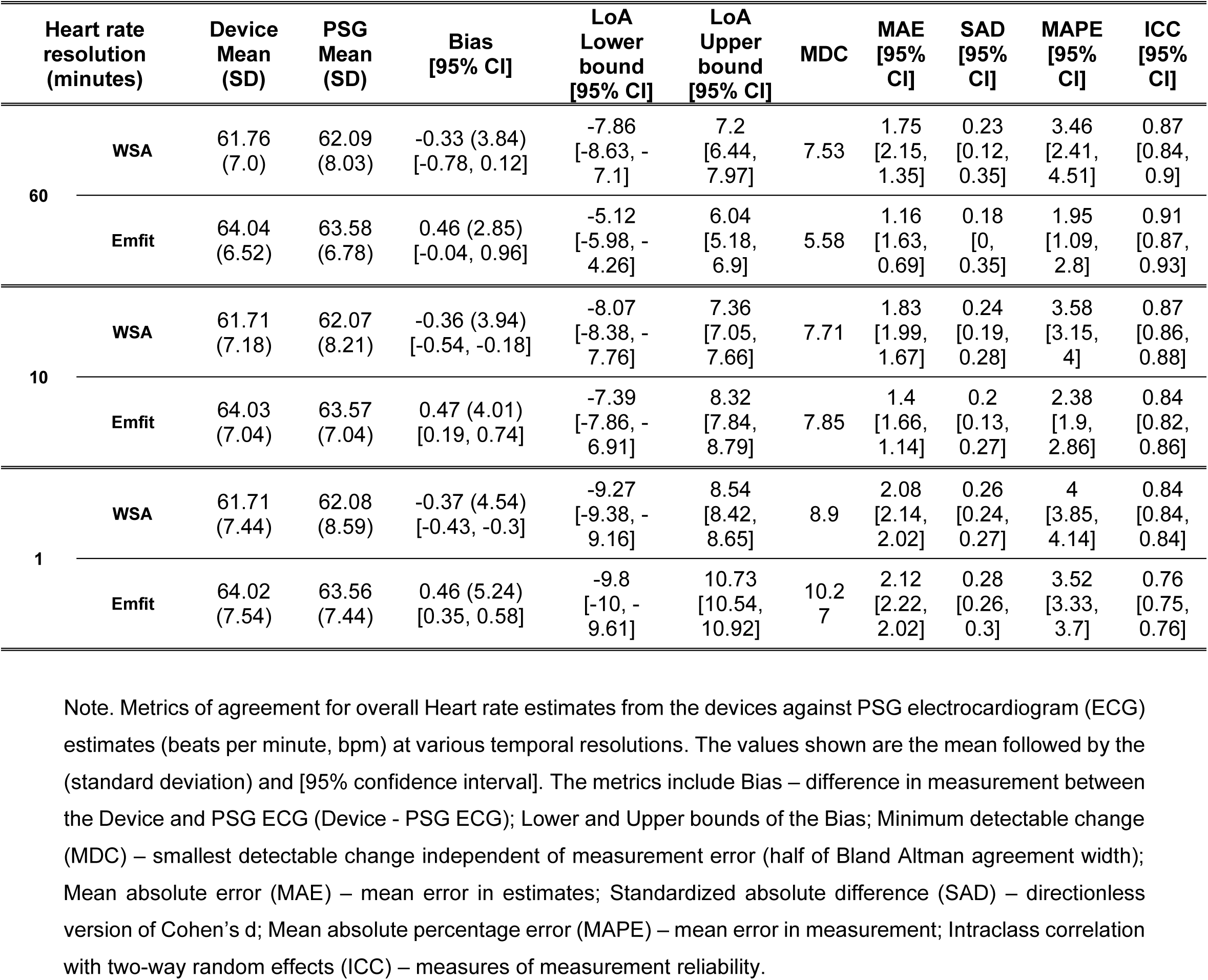
Effect of temporal resolution on reliability of estimates of heart rate.

**Table 3.** Effect of temporal resolution on reliability of estimates of breathing rate.

| Breathing rate resolution (minutes) |  | Device Mean (SD) | PSG Mean (SD) | Bias [95% CI] | LoA Lower bound [95% CI] | LoA Upper bound [95% CI] | MDC | MAE [95% CI] | SAD [95% CI] | MAPE [95% CI] | ICC [95% CI] |
| --- | --- | --- | --- | --- | --- | --- | --- | --- | --- | --- | --- |
| 60 | WSA | 13.67 (1.51) | 14.58 (1.87) | -0.91 (1.28)<br>[-1.06, -0.76] | -3.42<br>[-3.67, -3.17] | 1.6<br>[1.34, 1.85] | 2.51 | 1<br>[1.14, 0.86] | 0.59<br>[0.47, 0.71] | 6.48<br>[5.73, 7.22] | 0.72<br>[0.65, 0.77] |
|  | Emfit | 14.18 (1.86) | 14.33 (1.85) | -0.14 (1.66)<br>[-0.44, 0.15] | -3.39<br>[-3.9, -2.89] | 3.11<br>[2.6, 3.62] | 3.25 | 0.92<br>[1.16, 0.67] | 0.5<br>[0.32, 0.67] | 6.35<br>[4.68, 8.02] | 0.6<br>[0.47, 0.7] |
|  | Somnofy | 13.89 (1.65) | 14.31 (1.8) | -0.42 (1.19)<br>[-0.65, -0.19] | -2.76<br>[-3.16, -2.36] | 1.92<br>[1.52, 2.31] | 2.34 | 0.55<br>[0.77, 0.33] | 0.32<br>[0.13, 0.51] | 3.47<br>[2.27, 4.67] | 0.76<br>[0.67, 0.83] |
| 10 | WSA | 13.67 (1.67) | 14.6 (2.2) | -0.93 (1.67)<br>[-1.01, -0.86] | -4.21<br>[-4.34, -4.08] | 2.34<br>[2.21, 2.47] | 3.28 | 1.14<br>[1.21, 1.07] | 0.59<br>[0.54, 0.63] | 7.29<br>[6.94, 7.65] | 0.63<br>[0.61, 0.66] |
|  | Emfit | 14.16 (2.2) | 14.35 (2.17) | -0.18 (2.1)<br>[-0.33, -0.04] | -4.31<br>[-4.56, -4.06] | 3.94<br>[3.69, 4.19] | 4.12 | 1.09<br>[1.22, 0.96] | 0.5<br>[0.43, 0.57] | 7.42<br>[6.6, 8.25] | 0.54<br>[0.48, 0.58] |
|  | Somnofy | 13.87 (1.77) | 14.35 (2.05) | -0.48 (1.49)<br>[-0.59, -0.36] | -3.41<br>[-3.6, -3.21] | 2.45<br>[2.26, 2.65] | 2.93 | 0.68<br>[0.79, 0.57] | 0.36<br>[0.28, 0.43] | 4.2<br>[3.64, 4.76] | 0.69<br>[0.65, 0.73] |
| 1 | WSA | 13.67 (2.04) | 14.61 (2.61) | -0.93 (2.32)<br>[-0.97, -0.9] | -5.48<br>[-5.54, -5.42] | 3.61<br>[3.56, 3.67] | 4.55 | 1.51<br>[1.54, 1.48] | 0.64<br>[0.63, 0.66] | 9.85<br>[9.68, 10.02] | 0.51<br>[0.5, 0.52] |
|  | Emfit | 14.17 (2.5) | 14.35 (2.57) | -0.18 (2.73)<br>[-0.24, -0.12] | -5.54<br>[-5.65, -5.44] | 5.18<br>[5.07, 5.28] | 5.36 | 1.6<br>[1.65, 1.56] | 0.63<br>[0.61, 0.65] | 11.07<br>[10.74, 11.4] | 0.42<br>[0.4, 0.44] |
|  | Somnofy | 13.87 (1.91) | 14.35 (2.43) | -0.47 (1.99)<br>[-0.52, -0.43] | -4.38<br>[-4.46, -4.3] | 3.43<br>[3.35, 3.51] | 3.9 | 1.06<br>[1.1, 1.01] | 0.48<br>[0.46, 0.51] | 6.76<br>[6.54, 6.99] | 0.58<br>[0.57, 0.6] |

Note. Metrics of agreement for overall Heart rate PSG respiratory inductance plethysmography thorax (RIP Thorax) estimates (cycles per minute [cpm]). The values shown are the mean followed by the (standard deviation) and [95% confidence interval]. The metrics include Bias – difference in measurement between the Device and PSG ECG (Device - PSG ECG); Lower and Upper bounds of the Bias; Minimum detectable change (MDC) – smallest detectable change independent of measurement error (half of Bland Altman agreement width); Mean absolute error (MAE) – mean error in estimates; Standardized absolute difference (SAD) – directionless version of Cohen's d Mean absolute percentage error (MAPE) – mean error in measurement; Intraclass correlation with two-way random effects (ICC) – measures of measurement reliability.

For all devices and both heart and breathing rate the CDFs become steeper with increasing duration of the time window over which these variables computed. For both heart rate and breathing rate, the agreement (measured by ICC) with the PSG reference estimates increased with decreasing temporal resolution (Supplemental Figure 3). On closer inspection of the CDFs, we find that for the heart rate estimates, the error at the 90^th^ percentile is lower for WSA compared to Emfit for the 1 minute and 60 minute estimations (overall error < 4 bpm). For the Emfit, the median (50^th^ percentile) error of the heart rate estimates became smaller with increasing duration of the time window, but the WSA median error was always close to 1. For breathing rate, at all three resolutions, 50 % of the estimates had an error < 1 cpm. When we inspected the 90th percentile error at 1 minute, we found that Somnofy had the lowest error (2.56 cpm) followed by WSA (3.46 cpm) and Emfit (4.26 cpm). This trend was seen for lower resolutions as well. At the 50^th^ percentile we see the effect of discrete breathing rate output from WSA on the 50^th^ percentile error, where the error of the other two devices falls below 0.5 while the WSA error does not. The detailed discussion of the effects of temporal resolution on the agreement between the device and PSG vital signs estimate is provided in the supplemental materials.

### Estimating breathing disturbances during sleep

#### WSA Snore

The results of the WSA snore analysis and an example of the overnight time course of the snore data are depicted in Figure 6 and Supplementary Figure 6. Out of the 35 participants, snore data from both PSG and WSA were available for 30 participants (PSG snore sensor data were not available for two and WSA snore data were unavailable for three participants). Of the remaining 30 participants, it was determined from PSG snore sensor data that eight participants did not have any form of snoring whereas 22 participants snored. The WSA incorrectly determined five of these 22 participants (16.7 %) to have no snoring leading to a moderate performance with a Matthew’s CC of 0.69 (see Figure 6 c).

**Figure 6.**
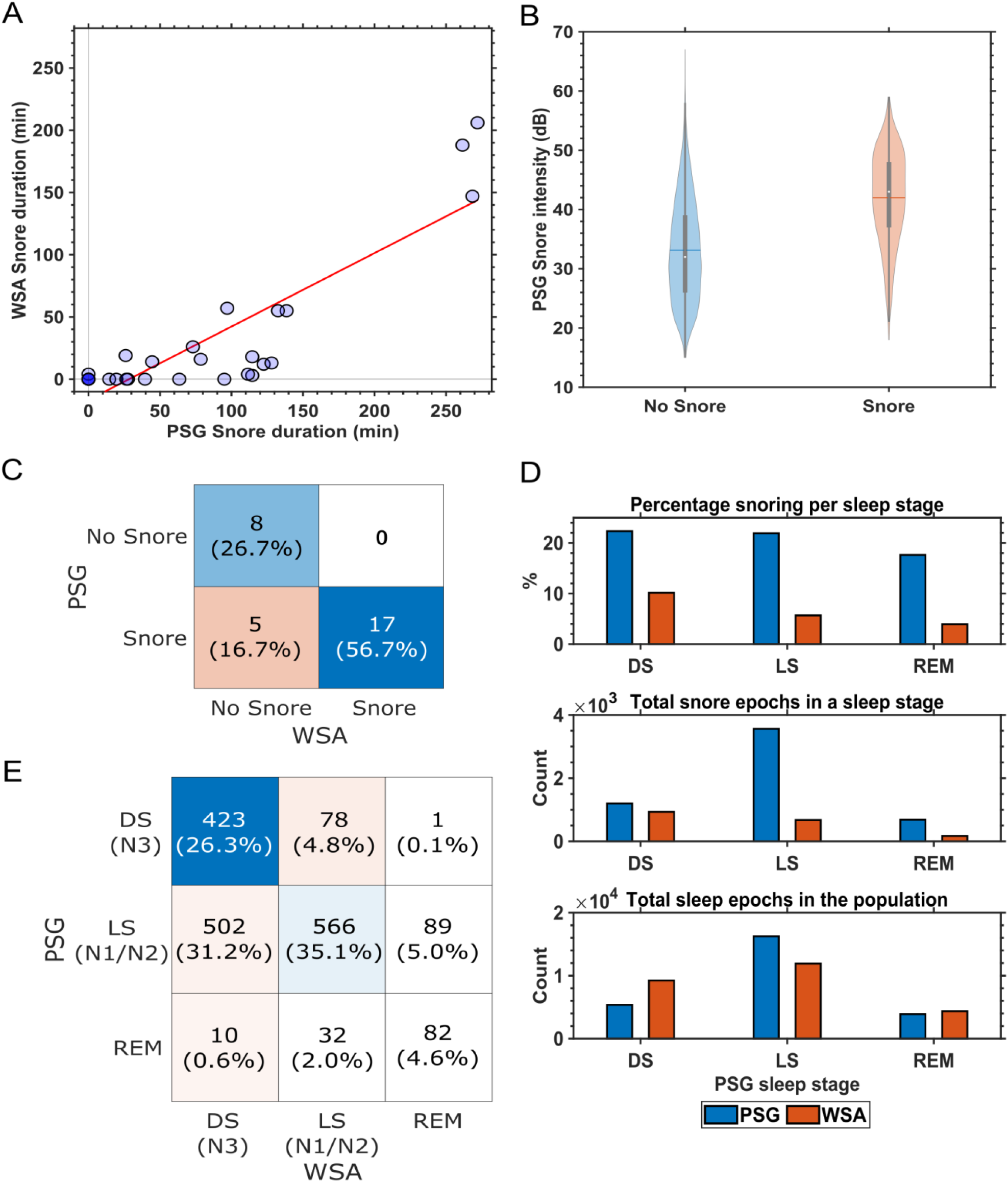
WSA Snore analysis. **A**. Concordance of PSG assessed snore duration and Withings sleep analyser (WSA) snore (n=30). The linear fit is depicted by the red line. **B.** Snore Intensity as detected by the PSG snore sensor during WSA predicted snore and no snore, **C.** Confusion matrix for participants identified as snorer of no snorer (n=30), **D.** Distribution of snore events across PSG derived sleep stages for both PSG and WSA, **E.** Confusion matrix of epoch-to-epoch (EBE) concordance between PSG and WSA during WSA snore events (Total WSA snore epochs: 1611).

**Figure 7.**
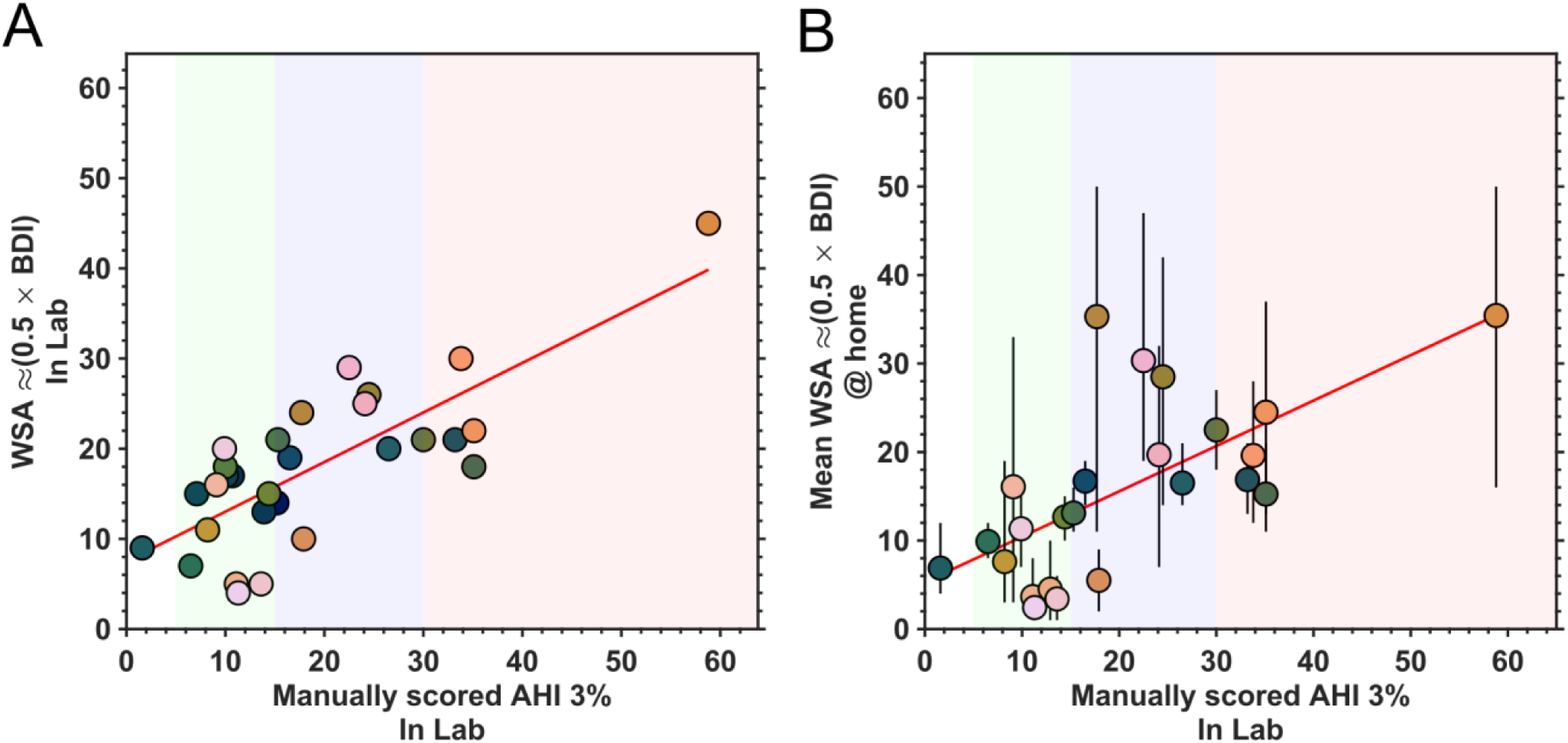
Relationship between polysomnography (PSG) based apnea-hypopnea index (AHI) and Withings sleep analyser (WSA) breathing disturbance index (BDI). **A**. in-laboratory (n=35) **B.** at home (n=29). The linear fit is depicted by the red line. AHI reference ranges are: No apnea – 0 to 4, Mild (green) – 5 to 14, Moderate – 15 to 29 (blue) and Severe – ≥30 (red). For the WSA AHI estimates at home, each data point depicts the mean per participant, and the vertical bars depict the minimum and maximum values.

PSG determined that the 22 snorers had a nightly snoring duration ranging between 10 to 270 minutes (see Figure 6 a). The concordance between the snore duration estimates between the WSA and PSG was high (r^2^ = 0.76, p < 0.001, n = 30). When the snoring intensity determined by the PSG snore sensor was grouped based on the WSA snore labels (Figure 6 b), we found that on average the intensity of the PSG detected snore events that were not detected by WSA was lower than the intensity of the snore events detected by both PSG and WSA, but there was a considerable overlap between the distributions.

Overall snore events were underestimated by the WSA compared to PSG. The distribution of snoring events as determined by the PSG snore sensor showed that snoring was present across all sleep states with > 20 % of all NREM epochs having snore events while for REM epochs this was 17.48 % (see Figure 6 d). On the other hand, when the distribution of snore events automatically identified by WSA was analysed the WSA determined snore events were high during LS and DS and low during REM (see Figure 6 e). The corresponding confusion matrix between the PSG consensus sleep stage and Withings sleep stage prediction during WSA identified snore events is depicted in Figure 6 e which shows that the WSA doesn’t score wake when snore events are detected. WSA also scored more snore epochs as deep sleep followed by light sleep and rapid eye movement sleep.

#### WSA apnea-hypopnea index

WSA summary measures relevant to breathing disorders are the breathing disorder index (BDI) and apnea-hypopnea index (AHI). WSA BDI was available for 29 participants in the laboratory and 24 participants at-home (a total of 222 nights available). Both WSA AHI and BDI were available for only 64 nights across 7 participants at home. Upon inspection of potential correlations between the WSA AHI and WSA BDI, we found that the WSA BDI was double the value of WSA AHI (WSA BDI =(2.006×WSA AHI) – 0.133; r^2^ = 0.999, p < 0.001, n = 64; see Supplementary Figure 7). Using this inference, we used half BDI as the proxy for WSA AHI for the remainder of the analysis.

We found a high correlation between the WSA AHI and PSG AHI in the laboratory (r^2^ = 0.59, p < 0.001, n = 29, Figure 6a). The MAE was 6.49 [4.89, 8.10] events/hour. We further explored the relation between the PSG AHI in the laboratory and mean WSA AHI at-home (see Figure 6 b). We found that there was moderate level of correction between the two (r^2^ = 0.44, p<0.001, n = 24). There was also high level of agreement between the WSA AHI one night prior to the laboratory and both in-laboratory WSA AHI (r^2^ = 0.74, p < 0.001, n = 15) and PSG AHI (r^2^ = 0.59, p < 0.001, n = 16). We have depicted the WSA AHI (half WSA BDI) for the 24 participants data available at-home as a heatmap showing the night-to-night variability and missing values in Supplementary Figure 8.

## Discussion

### Principal Findings

In this study, we provide an evaluation of three contactless technologies for monitoring heart rate, breathing rate and breathing disturbance during sleep in older men and women. Overall, the contactless technologies provided heart rate (WSA and Emfit) and breathing rate (all three devices) estimates with acceptable agreement compared to standard reference estimates from PSG ECG and RIP thorax in this older population. We also found that these devices can be used for detecting respiratory events including apnea and snoring in this population of older men and women with stable comorbidities.

We were able to successfully collect data at-home with limited (< 5 %) data loss. The data loss was primarily due to Wi-Fi dropouts where the device spontaneously lost connection to the Wi-Fi network. Overall, this demonstrates the ability of these contactless devices to reliable collect continuous vital signs data remotely in the community with little oversight and maintenance.

The devices also captured the time course of vital signs during sleep in good agreement with PSG and with relatively small differences in performance between the devices. The heart rate estimate range of both WSA and Emfit were narrower than the AAMI recommended minimum allowable range of 30 to 200 bpm, with WSA having a more limited range compared to Emfit [48]. WSA performed somewhat better than Emfit at capturing heart rate trends while for the breathing rate, Somnofy performed the best followed by WSA and Emfit. The contactless technologies provided estimates of heart rate and breathing which when averaged across the night, were in very good agreement with the PSG estimates. Outliers in both breathing rate and heart rate agreement plots were found to be originating from participants with severe breathing disturbances or significant abnormal cardiac rhythm. That the overall agreement between contactless technologies derived estimates and estimates derived from PSG becomes poorer when the time period over which the estimate is computed becomes shorter is not surprising but puts limitations on the use-cases in which these devices can be applied. Improvement in the estimates from WSA with reducing temporal resolution was limited by the discretised or rounded output of vital signs from the WSA with 50% of the estimates having a minimum error of 1 bpm.

At 1 minute resolution, the under-mattress devices had an acceptable accuracy with MAE < 2.12 bpm and MAPE < 5 % for heart rate estimate which is lower than the errors reported in the literature for wearable technologies during daily activities and for many contactless technologies during sleep [24, 28]. The breathing rate estimates at 1 minute resolution were acceptable across all three devices with a MAE ≤ 1.6 cpm and MAPE > 12 % and was comparable to other contactless technologies in in previous evaluations in young participants [28]. The accuracy of both the vital signs estimates was higher during sleep than wake primarily due to reduced body movements. Somnofy, the best performing device in estimating breathing rate was also the best performing device in our evaluation of sleep stage classification performance [36].

Although in our evaluation in older participants, all the compared contactless devices provided acceptable performance, this performance was poorer than previously reported in studies (Emfit and Somnofy) in a younger population. In the evaluation conducted by Ranta et al., in a population of 34 participants with a median age of 32 years, the Emfit had an MAE of 1.34 bpm for heart rate and a MAE of 0.59 cpm for breathing rate [33]. In contrast, in the evaluation conducted by Toften et al, Somnofy had an MAE of 0.18 cpm in a population of 37 participants with a mean age of 32.6 years [32]. Although the vital signs estimates of WSA have been used in large scale studies, there is no existing evaluation of WSA estimated vital signs in the literature to the best of our knowledge [52, 53].

The higher accuracy of Somnofy in estimating breathing rate compared to the under-mattress devices can be attributed to the device not estimating breathing rate when the signal quality is affected by body movements. Although this leads to some loss of data, this also leads to better accuracy depicting the need for a signal quality index associated with the device generated vital signs estimates.

Finally, our evaluation of revealed that the WSA snore and BDI estimates were accurate and the performance of the WSA AHI in terms of MAE in our study (MAE: 6.49 events/ hours; N=29) was better than the results reported in Edouard et al (MAE: 9.5 events/ hours; N=118; mean age = 49.3 years) [54]. To the best of our knowledge, the WSA snore has not been previously evaluated in other studies. The breathings disturbance detection has been performed using Emfit raw ballistography data in literature, but these algorithms are not open source or available directly from the manufacturer and hence not used in our analysis [55, 56]. The availability of these breathing disturbance estimates along with the acceptable agreement of the vital signs measures generated by the contactless technologies demonstrates their immediate potential usefulness in population wide deployment for home monitoring and care [57].

### Limitations

One of the limitations of the work is that the data synchronization of the PSG reference data and the device data is based on best alignment of the vital signs data and hypnogram which is not ideal. Secondly, the algorithms used by the different devices in deriving the ballistography signal and heart rate and breathing rate information is hidden due to their proprietary nature and hence the interpretability of several observations made in this study such as bounded nature of the output vital signs, outliers and unavailability of data is limited.

### Conclusion and outlook

With their ability to reliably collect heart rate, breathing rate and breathing disturbance data longitudinally and at scale, contactless technologies have the potential to be a powerful tool for unintrusive remote vital signs monitoring in community dwelling older adults and PLWD populations. Applications range from early detection of abnormalities and deterioration of health to monitoring the impact of interventions to improve health, i.e., treatment of sleep apnea; together these applications could improve overall home care. They also allow investigation into the night-to-night variation in sleep and vital signs and how this variation associates with health outcomes and daytime function. Such approaches have already shown that night to night variation in sleep apnea associates with uncontrolled hypertension [57] and that night-to-night variation in sleep continuity associated with day-to-day variation in symptoms in people living with Alzheimer’s [58].

## Acknowledgement

This work was supported by the UK Dementia Research Institute, Care Research & Technology Centre at Imperial College, London and the University of Surrey, Guildford, United Kingdom which receives its funding from UK DRI Ltd, funded by the UK Medical Research Council, Alzheimer’s Society and Alzheimer’s Research UK. We thank staff members of the Clinical Research Facility for their help with data collection and members of Care Research & Technology for helpful discussions.

KR conducted the data exploration and analysis and prepared the manuscript. DJD conceived the study and contributed to writing of the manuscript. CdM, GA, HH and VR contributed to the design of the study, were responsible for participant recruitment and screening, and study conduct. The devices were setup and datasets were downloaded and curated by DL, GA, KR, CdM and VR. All the authors contributed to the data collection and contributed to finalising the manuscript.

## Data Availability

The data used in this study are available from the co-author Ciro della Monica upon reasonable request. Contact.

