## Supplemental Materials for "Reliable Contactless Monitoring of Heart Rate, Breathing Rate and Breathing Disturbance During Sleep in Aging: A Digital Health Technology Evaluation Study"

Name: Dr Kiran Kumar Guruswamy Ravindran

Address: Surrey Sleep Research Centre, University of Surrey, GU2 7XP

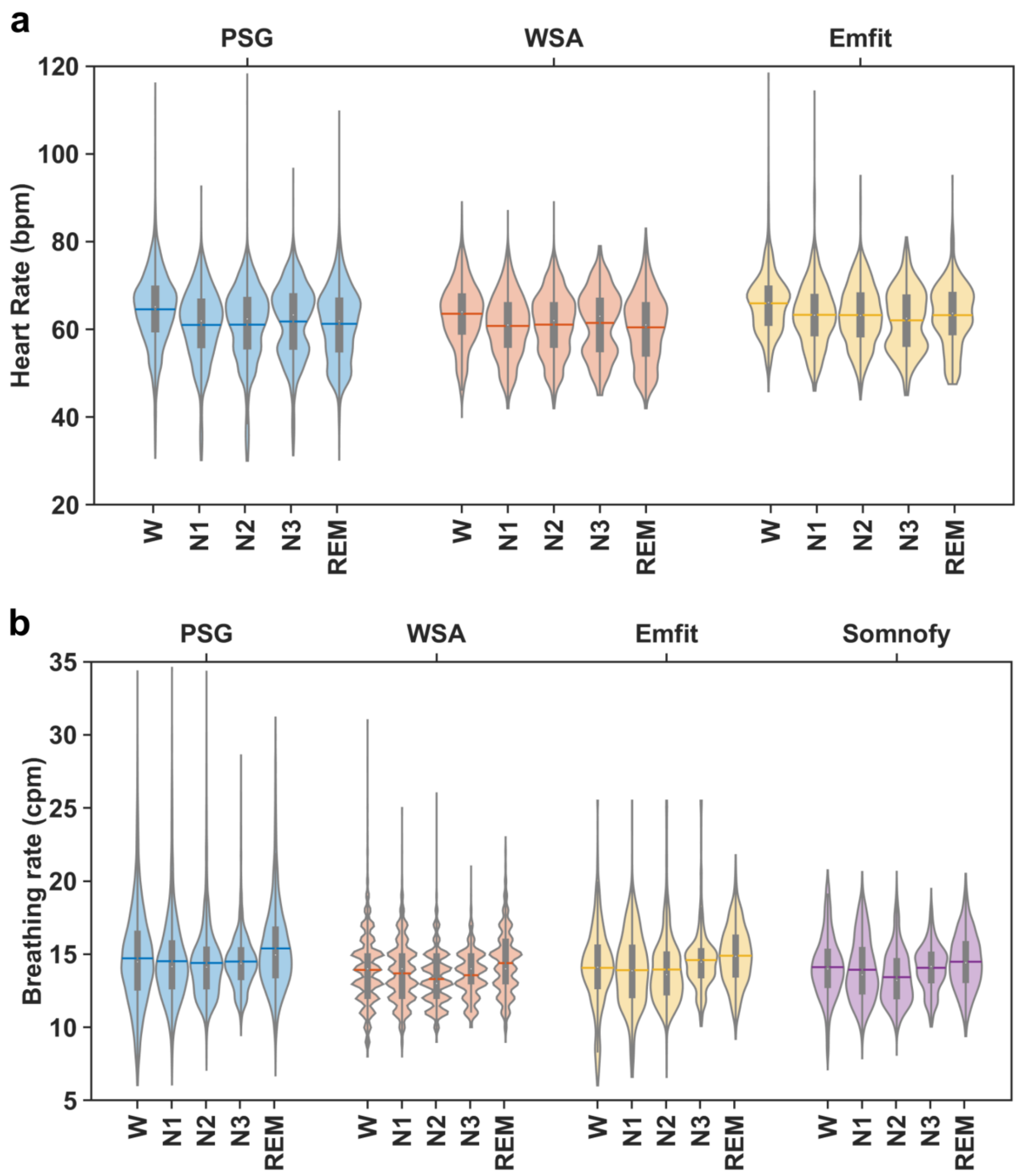


**Supplemental Figure 1. Vitals signs distribution across different sleep states in the laboratory.** For all devices the consensus PSG hypnogram was used to select the data Labels at the bottom horizontal axis denotes the sleep stage while the devices names are indicated on the top. Each violin represents the distribution of heart rate (top panel) and breathing rate (bottom panel) estimates. The boxplot at the centre of the violin depicts the median (circle at the centre), 1st and 3rd quartiles (lower and upper ends of the box) and the whiskers depict the 1.5 × IQR (inter quartile range). The solid-coloured line in the violins depict the mean.

**Supplementary Table 1. Sleep stage specific all night heart rate concordance**

| Heart rate (beats per min.) | Device Mean (SD) | PSG Mean (SD) | Bias [95% CI] | LoA Lower bound [95% CI] | LoA Upper bound [95% CI] | MDC | MAE [95% CI] | SAD [95% CI] | MAPE [95% CI] | ICC [95% CI] |
|---|---|---|---|---|---|---|---|---|---|---|
| **Withings sleep analyser (WSA)** | | | | | | | | | | |
| **LS** | 60.81 (6.65) | 61.1 (7.79) | -0.29 (3.95) [-1.64 1.07] | -8.02 [-10.36 - 5.68] | 7.44 [5.11 9.78] | 7.73 | 1.59 [2.83 0.35] | 0.22 [-0.12 0.56] | 3.3 [-0.12 6.73] | 0.85 [0.73 0.92] |
| **DS** | 61.58 (7.09) | 61.88 (8.05) | -0.3 (3.31) [-1.43 0.84] | -6.78 [-8.74 - 4.82] | 6.18 [4.22 8.14] | 6.48 | 1.46 [2.48 0.44] | 0.19 [-0.14 0.53] | 2.88 [0.19 5.57] | 0.91 [0.82 0.95] |
| **REM** | 61.68 (6.9) | 62.25 (7.57) | -0.58 (2.68) [-1.5 0.34] | -5.82 [-7.41 - 4.23] | 4.67 [3.08 6.25] | 5.24 | 1.41 [2.22 0.61] | 0.2 [-0.14 0.54] | 2.56 [0.7 4.43] | 0.93 [0.87 0.96] |
| **NREM** | 60.99 (6.64) | 61.27 (7.77) | -0.28 (3.82) [-1.59 1.03] | -7.77 [-10.04 - 5.51] | 7.21 [4.94 9.48] | 7.49 | 1.53 [2.74 0.33] | 0.22 [-0.12 0.55] | 3.17 [-0.12 6.47] | 0.86 [0.74 0.93] |
| **Wake** | 63.24 (6.49) | 64.3 (7) | -1.06 (3.21) [-2.17 0.04] | -7.36 [-9.26 - 5.46] | 5.23 [3.33 7.13] | 6.3 | 2.15 [3.04 1.26] | 0.32 [-0.01 0.66] | 3.69 [1.73 5.65] | 0.89 [0.79 0.94] |
| **EMFIT** | | | | | | | | | | |
| **LS** | 63.11 (5.82) | 62.59 (6.72) | 0.52 (1.36) [-0.21 1.25] | -2.15 [-3.42 - 0.88] | 3.19 [1.92 4.46] | 2.67 | 0.86 [1.48 0.25] | 0.14 [-0.37 0.66] | 1.52 [0.3 2.75] | 0.98 [0.93 0.99] |
| **DS** | 62.17 (7.34) | 62.28 (7.61) | -0.12 (0.92) [-0.61 0.37] | -1.92 [-2.78 - 1.07] | 1.69 [0.83 2.54] | 1.8 | 0.56 [0.94 0.17] | 0.08 [-0.44 0.59] | 0.86 [0.25 1.47] | 0.99 [0.98 1] |
| **REM** | 63.48 (6.25) | 63.52 (6.82) | -0.03 (1.18) [-0.66 0.6] | -2.35 [-3.46 - 1.25] | 2.29 [1.19 3.39] | 2.32 | 0.63 [1.16 0.11] | 0.1 [-0.42 0.62] | 1.06 [0.07 2.05] | 0.98 [0.95 0.99] |

| | | | | | | | | | | |
|---|---|---|---|---|---|---|---|---|---|---|
| **NREM** | 62.89 (6.06) | 62.51 (6.84) | 0.38 (1.21) [-0.26 1.03] | -1.99 [-3.12 - 0.87] | 2.76 [1.63 3.88] | 2.37 | 0.77 [1.3 0.24] | 0.12 [-0.39 0.64] | 1.34 [0.3 2.38] | 0.98 [0.95 0.99] |
| **Wake** | 65.76 (5.22) | 65.34 (6) | 0.42 (3.31) [-1.35 2.18] | -6.08 [-9.16 - 2.99] | 6.91 [3.82 9.99] | 6.49 | 2.02 [3.41 0.64] | 0.37 [-0.14 0.89] | 3.32 [0.8 5.84] | 0.83 [0.57 0.94] |

Note. Metrics of agreement for overall heart rate of the devices and the estimates during different vigilance states (light sleep (LS), deep sleep (DS), rapid eye movement sleep (REM) and wake) against PSG electrocardiogram (ECG) estimates (beats per minute, bpm). The values shown are the mean followed by the (standard deviation) and [95% confidence interval]. The metrics include Bias – difference in measurement between the Device and PSG ECG (Device - PSG ECG); Lower and Upper bounds of the Bias; Minimum detectable change (MDC) – smallest detectable change independent of measurement error (half of Bland Altman agreement width); Mean absolute error (MAE) – mean error in estimates; Standardized absolute difference (SAD) – directionless version of Cohen’s d; Mean absolute percentage error (MAPE) – mean error in measurement; Intraclass correlation with two-way random effects (ICC) – measure of measurement reliability.

**Supplementary Table 2. Sleep stage specific all night breathing rate concordance**

| Breathing rate (cycles per min.) | Device Mean (SD) | PSG Mean (SD) | Bias [95% CI] | LoA Lower bound [95% CI] | LoA Upper bound [95% CI] | MDC | MAE [95% CI] | SAD [95% CI] | MAPE [95% CI] | ICC [95% CI] |
|---|---|---|---|---|---|---|---|---|---|---|
| Withings sleep analyser (WSA) | | | | | | | | | | |
| **LS** | 13.44 (1.53) | 14.46 (1.8) | -1.02 (1.11) [-1.4 -0.64] | -3.19 [-3.84 - 2.53] | 1.15 [0.49 1.81] | 2.17 | 1.02 [1.4 0.64] | 0.62 [0.28 0.96] | 6.75 [4.72 8.77] | 0.78 [0.61 0.88] |
| **DS** | 13.45 (1.48) | 14.42 (1.51) | -0.97 (0.77) [-1.24 - 0.71] | -2.49 [-2.94 - 2.03] | 0.54 [0.08 1] | 1.51 | 0.97 [1.24 0.71] | 0.66 [0.32 1] | 6.67 [5.06 8.29] | 0.87 [0.75 0.93] |
| **REM** | 14.39 (1.64) | 15.3 (1.95) | -0.91 (1.3) [-1.36 - 0.47] | -3.46 [-4.23 - 2.69] | 1.63 [0.86 2.4] | 2.54 | 0.99 [1.41 0.56] | 0.55 [0.22 0.89] | 6.09 [3.83 8.34] | 0.74 [0.54 0.86] |
| **NREM** | 13.44 (1.5) | 14.44 (1.72) | -1 (0.99) [-1.34 - 0.66] | -2.95 [-3.53 - 2.36] | 0.94 [0.36 1.53] | 1.95 | 1 [1.34 0.66] | 0.63 [0.29 0.97] | 6.69 [4.83 8.56] | 0.81 [0.66 0.9] |
| **Wake** | 13.87 (1.35) | 14.74 (2.05) | -0.87 (1.65) [-1.44 -0.3] | -4.11 [-5.09 - 3.13] | 2.37 [1.39 3.35] | 3.24 | 1.29 [1.75 0.82] | 0.75 [0.41 1.09] | 8.43 [5.72 11.14] | 0.55 [0.27 0.74] |
| **Emfit** | | | | | | | | | | |
| **LS** | 14.04 (1.67) | 14.21 (1.7) | -0.17 (1.18) [-0.8 0.47] | -2.49 [-3.59 - 1.39] | 2.16 [1.05 3.26] | 2.32 | 0.69 [1.2 0.18] | 0.42 [-0.09 0.94] | 4.81 [1.46 8.16] | 0.75 [0.43 0.91] |
| **DS** | 14.29 (1.48) | 14.36 (1.71) | -0.07 (1.16) [-0.69 0.54] | -2.34 [-3.41 - 1.26] | 2.19 [1.12 3.27] | 2.27 | 0.66 [1.16 0.16] | 0.43 [-0.09 0.94] | 4.52 [1.26 7.77] | 0.74 [0.4 0.9] |
| **REM** | 14.89 (1.47) | 14.98 (1.4) | -0.09 (0.77) [-0.5 0.32] | -1.6 [-2.32 - 0.88] | 1.42 [0.71 2.14] | 1.51 | 0.48 [0.8 0.15] | 0.34 [-0.17 0.86] | 3.21 [1.08 5.34] | 0.86 [0.64 0.95] |
| **NREM** | 14.1 (1.61) | 14.23 (1.68) | -0.13 (1.1) [-0.72 0.45] | -2.28 [-3.3 -1.26] | 2.02 [1 3.04] | 2.15 | 0.65 [1.12 0.18] | 0.41 [-0.11 0.92] | 4.56 [1.52 7.59] | 0.78 [0.47 0.92] |

| | | | | | | | | | | |
|---|---|---|---|---|---|---|---|---|---|---|
| **Wake** | 13.93 (1.96) | 14.4 (2.21) | -0.48 (1.7) [-1.38 0.43] | -3.8 [-5.38 - 2.22] | 2.85 [1.27 4.43] | 3.33 | 1.24 [1.89 0.59] | 0.61 [0.1 1.13] | 8.64 [4.37 12.91] | 0.67 [0.28 0.87] |
| **Somnofy** | | | | | | | | | | |
| **LS** | 13.65 (1.76) | 14.14 (1.67) | -0.49 (1.03) [-1.02 0.03] | -2.5 [-3.42 - 1.58] | 1.52 [0.6 2.44] | 2.01 | 0.56 [1.07 0.06] | 0.34 [-0.16 0.84] | 3.82 [0.61 7.02] | 0.82 [0.57 0.93] |
| **DS** | 13.77 (1.6) | 14.29 (1.69) | -0.51 (1.01) [-1.03 0.01] | -2.5 [-3.4 -1.59] | 1.47 [0.56 2.38] | 1.98 | 0.52 [1.03 0] | 0.32 [-0.18 0.82] | 3.37 [0.2 6.53] | 0.81 [0.55 0.93] |
| **REM** | 14.49 (1.67) | 14.87 (1.42) | -0.38 (1) [-0.9 0.13] | -2.35 [-3.25 - 1.45] | 1.58 [0.68 2.48] | 1.96 | 0.61 [1.06 0.16] | 0.41 [-0.09 0.9] | 4.14 [1.11 7.16] | 0.79 [0.51 0.92] |
| **NREM** | 13.68 (1.72) | 14.16 (1.66) | -0.48 (0.97) [-0.98 0.02] | -2.38 [-3.24 - 1.51] | 1.41 [0.55 2.28] | 1.89 | 0.53 [1.01 0.04] | 0.32 [-0.18 0.82] | 3.57 [0.51 6.63] | 0.84 [0.6 0.94] |
| **Wake** | 13.6 (1.92) | 14.32 (2.17) | -0.72 (1.69) [-1.59 0.15] | -4.04 [-5.55 - 2.52] | 2.59 [1.08 4.11] | 3.31 | 1.16 [1.89 0.44] | 0.59 [0.09 1.08] | 7.81 [3.4 12.22] | 0.66 [0.28 0.86] |

Note. Metrics of agreement for Overall breathing rate of the devices and the estimates during different vigilance states (light sleep (LS), deep sleep (DS), rapid eye movement sleep (REM) and wake) against PSG respiratory inductance plethysmography thorax (RIP Thorax) estimates (cycles per minute). The values shown are the mean followed by the (standard deviation) and [95% confidence interval]. The metrics include Bias – difference in measurement between the Device and PSG RIP Thorax (Device - PSG RIP Thorax); Lower and Upper bounds of the Bias; Minimum detectable change (MDC) – smallest detectable change independent of measurement error (half of Bland Altman agreement width); Mean absolute error (MAE) – mean error in estimates; Standardized absolute difference (SAD) – directionless version of Cohen’s d; Mean absolute percentage error (MAPE) – mean error in measurement; Intraclass correlation with two-way random effects (ICC) – measures of measurement reliability.

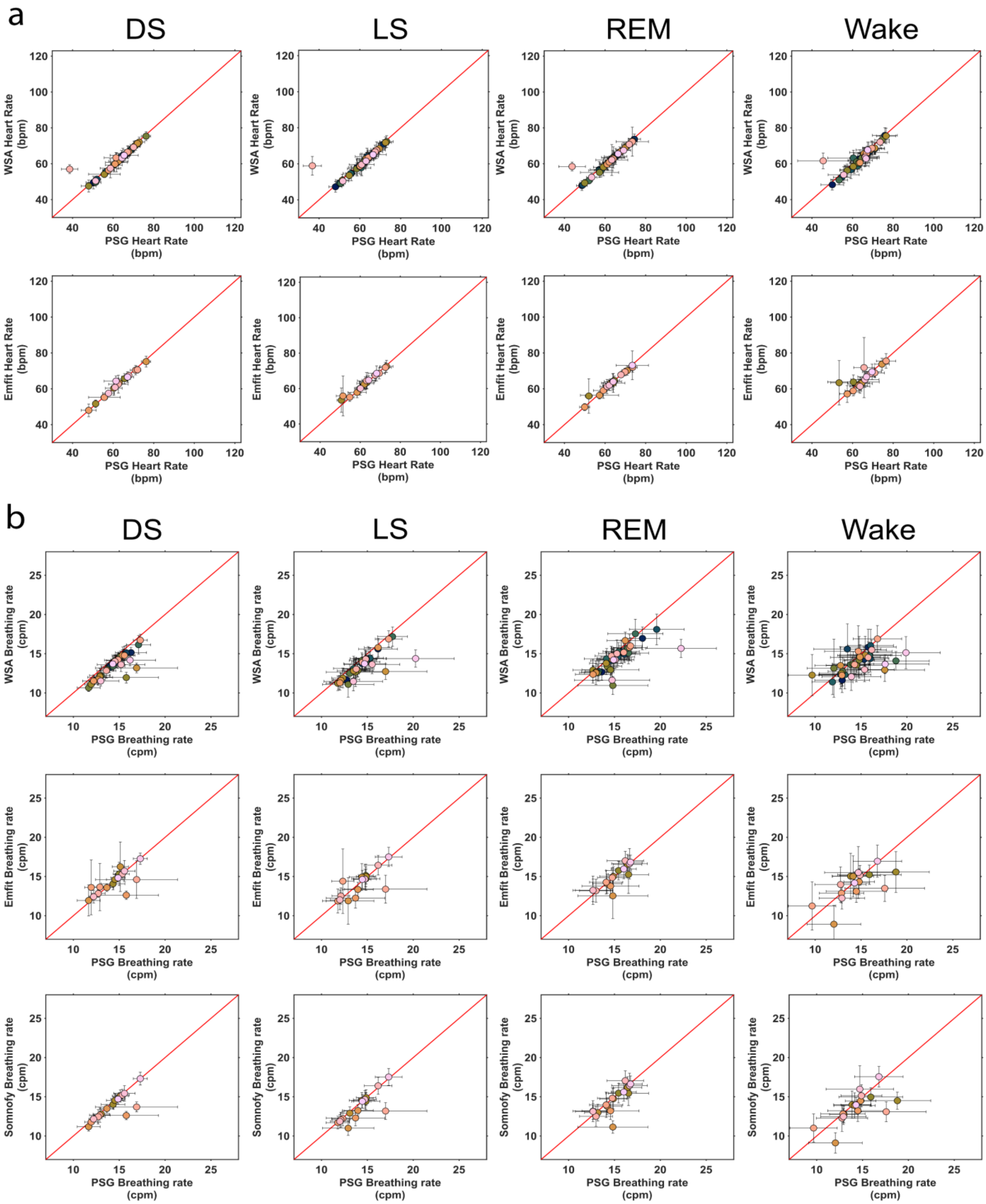


**Supplementary Figure 2. Association of vital signs recorded by the devices and PSG during different vigilance states. a.** Heart rate (beats per minute, bpm) **b.** Breathing rate (cycles per min, cpm). The four columns of the plots represent deep sleep (DS), light sleep (LS), rapid eye movement sleep (REM) and wake. The error bars represent the standard deviation of the estimate. The consensus polysomnography (PSG) was used to group sleep stage specific vital signs. The number of participants used in each of the devices are WSA [n=34], Emfit [n=16] and Somnofy [n=17].

### Vitals signs during different vigilance states

For both the contactless technologies and the PSG, the mean heart rates were highest during wake followed by REM and NREM. For the WSA, the concordance as measured by MAPE and ICC were the highest for REM followed by DS, LS, and wake. REM also had the lowest MDC (see Supplemental Table 1). For the Emfit, the heart rate concordance was the highest for DS, followed by REM, LS and wake and the MDC values follows the concordance estimates with DS having the lowest MDC while wake, the highest. For both the under-mattress devices, the SAD was the lowest for DS and highest for wake (Supplemental Figure 2 a).

The mean breathing rate was highest during REM. The DS mean breathing rate was higher than LS according to all three compared devices (see Supplemental table 2). The wake breathing rate on the other hand, was higher than NREM for WSA but lower than NREM for Emfit and Somnofy. The concordance to PSG was the highest for REM followed by DS, LS, and wake for the under-mattress devices while for Somnofy DS had the highest concordance followed by LS, REM, and wake. The MDC and SAD were the highest for wake across all devices. For Emfit and Somnofy, the wake MDC was followed by LS, DS and REM, but for the WSA wake was followed by REM, LS, and DS. SAD was highest for REM and lowest for DS in Somnofy but for the under-mattress devices REM had the lowest SAD. For WSA, LS had the highest SAD followed by DS while for Emfit DS SAD was higher than LS.

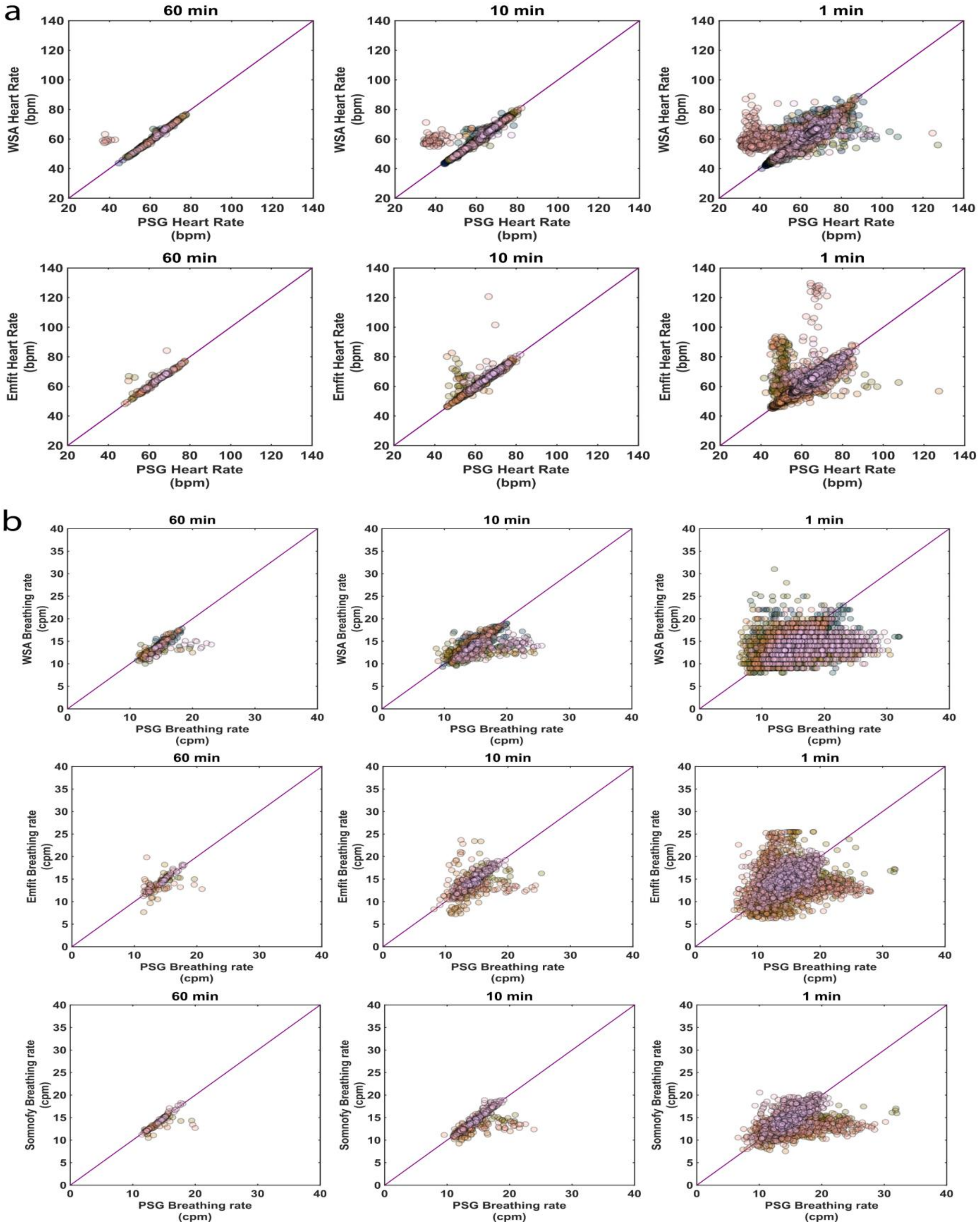


**Supplementary Figure 3. Effect of temporal resolution on accuracy of vital signs estimates. a.** Heart rate (WSA-top; Emfit-bottom) **b.** Breathing rate (WSA-top; Emfit-middle; Somnofy-bottom). The estimate resolutions from left to right are 60 min, 10 min and 1 min. The 45-degree line represents the line of unity i.e., perfect correlation. The number of participants (=nights) available for the devices are WSA [n=34], Emfit [n=16] and Somnofy [n=17].

### Effect of temporal resolution on the vital signs accuracy

In this section, we further discuss the effects of temporal resolution on the vital signs accuracy using standard Bland Altman measures. The results for heart rate and breathing rate comparisons are provided in supplemental figures 4 and 5 and table 3 and 4, respectively. It should be noted that the outliers in the Supplemental Figure 3, 4 and 5 correspond to participants with cardiac issues and severe breathing disturbance. These data represent a realistic measurement situation that occurs in the real world in older adult populations.

For WSA heart rate, the improvement in the accuracy of the estimates was higher for 10 minute estimates while it was low when compared between 10 minute and 60 minutes estimates. Accuracy of the Emfit heart rate estimates, on the other hand, continued to improve when the resolution was decreased to both 10 minutes and 60 minutes. The WSA heart rate had a higher agreement with the PSG reference estimates at 1 minute and 10 minute resolution while for the 60 minutes averages, Emfit heart rate agreement was higher.

The breathing rate estimates of all three contactless technologies showed marked improvement with decreasing resolution. At all three resolutions, the bedside radar, Somnofy, provided more accurate estimates compared to the under-mattress devices. Among the under-mattress devices, WSA had a better accuracy compared to Emfit. The same observation can be seen for MAPE and MDC while the estimate of dispersion, SAD, did not show a marked change between 10-minute and 60-minute estimates.

We further inspected the differences in the estimation errors between sleep and wake states. The sleep and wake CDFs along with a more detailed overview of the heart rate and breathing rate agreement with Bland-Altman plots at 1 minute resolution is provided in Supplementary Figure 4 and 5. The agreement measures for the same are provided in Supplementary Tables 3 and 4. We found that for both heart rate and breathing rate, the estimates during the wake epochs had considerably higher 90$^{th}$ percentile error compared to sleep. Emfit had the worst error in estimation during wake for both heart rate and breathing rate. For breathing rate, the Somnofy had the lowest error during wake.

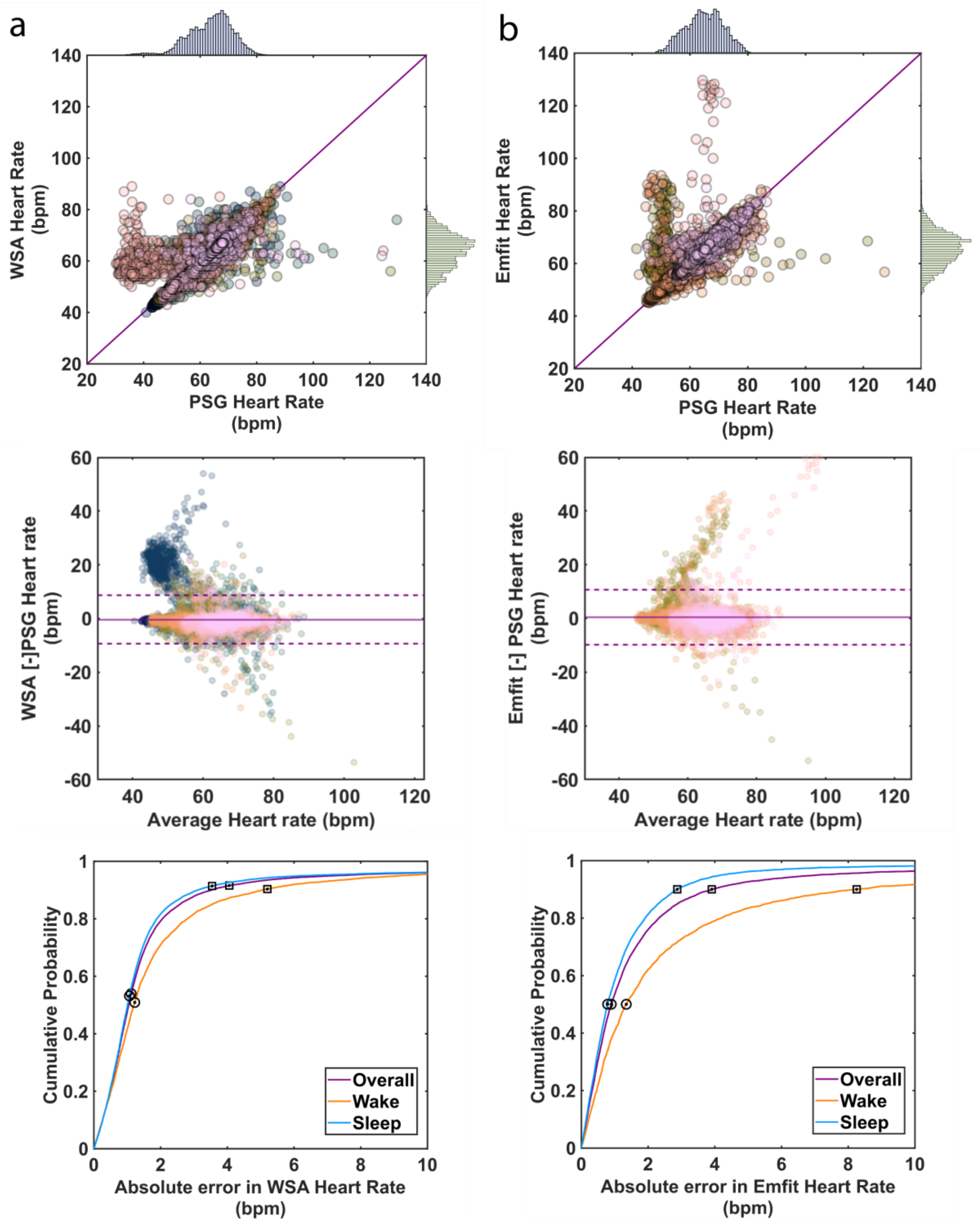


**Supplementary Figure 4. Evaluation of heart rate (beats per minute, bpm) estimates at one minute resolution. a** Withings sleep analyser (WSA) and **b** Emfit QS (Emfit). The top row depicts the scatter plots of PSG electrocardiogram vs Device heart rate along with the associated normalised heart rate histograms on the axis, the centre row depicts the Bland-Altman plots, and the bottom row depicts the cumulative density function of the absolute error of the heart rate estimates compared to PSG electrocardiogram estimates. The circled data point represents the median while the square represents the 90th percentile.

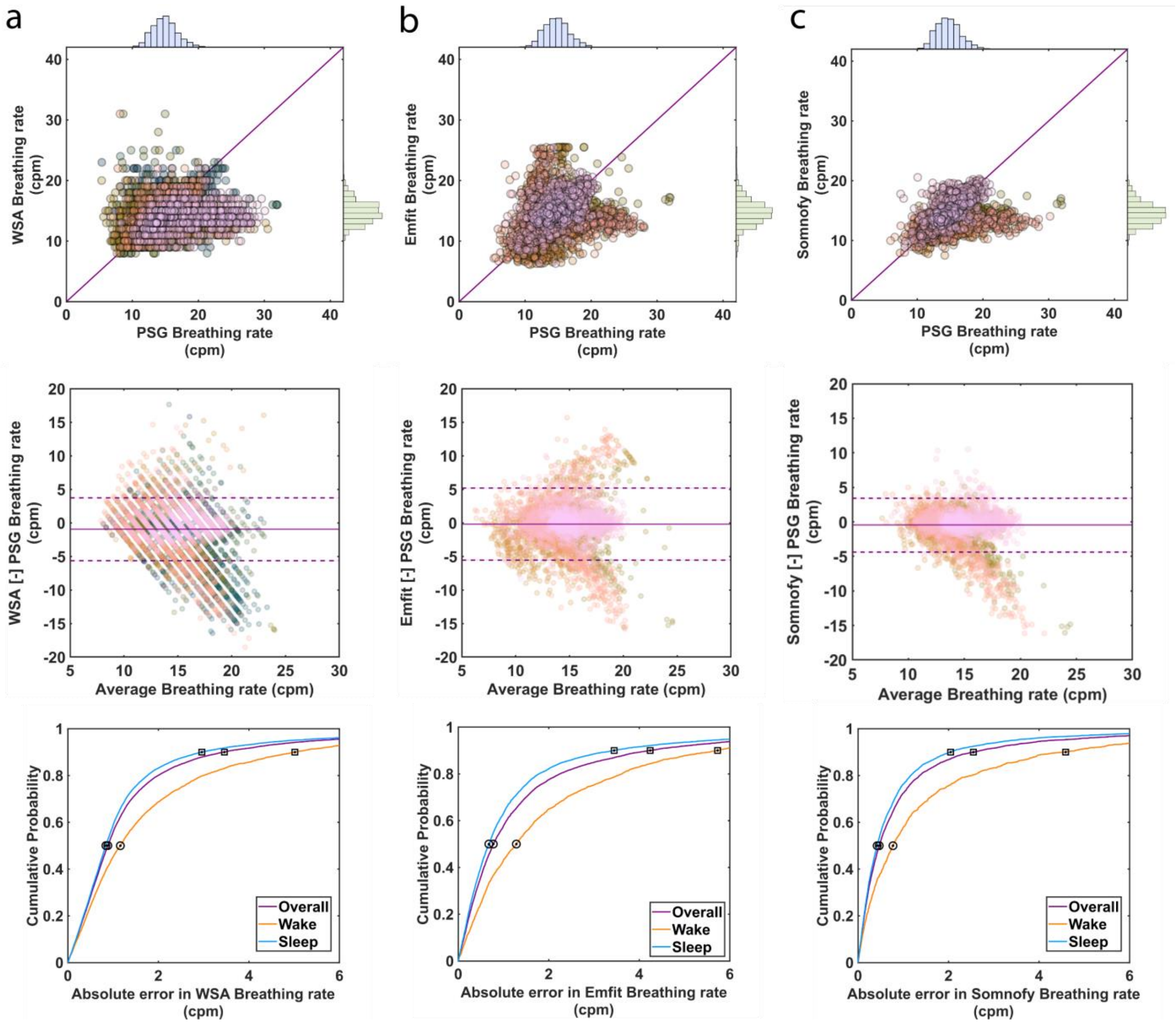


**Supplementary Figure 5. Evaluation of breathing rate (cycles per minute, cpm) estimates at one minute resolution. a** Withings sleep analyser (WSA) and **b** Emfit QS (Emfit). The top row depicts the scatter plots of PSG respiratory inductance plethysmography thorax (RIP Thorax) vs Device heart rate along with the associated normalised breathing rate histograms on the axis, the centre row depicts the Bland-Altman plots, and the bottom row depicts the cumulative density function of the absolute error of the breathing rate estimates compared to PSG RIP thorax estimates. The circled data point represents the median while the square represents the 90$^{th}$ percentile.

**Supplementary Table 3. Heart rate agreement measures in the laboratory (1 min)**

| Heart rate agreement | | Bias [95% CI] | LoA Lower bound [95% CI] | LoA Upper bound [95% CI] | MDC | SAD [95% CI] | MAPE [95% CI] | ICC [95% CI] |
|---|---|---|---|---|---|---|---|---|
| Overall | WSA | -0.38 (4.54) [-0.44, -0.31] | -9.28 [-9.39 -9.17] | 8.53 [8.41 8.64] | 8.9 | 0.26 [0.25 0.27] | 4 [3.85 4.15] | 0.76 [0.71 0.81] |
| | Emfit | 0.46 (5.24) [0.35 0.58] | -9.8 [-10 -9.61] | 10.73 [10.54 10.92] | 10.27 | 0.28 [0.26 0.3] | 3.52 [3.33 3.7] | 0.58 [0.48 0.68] |
| Sleep | WSA | -0.32 (4.52) [-0.39 -0.24] | -9.18 [-9.3 -9.05] | 8.55 [8.42 8.67] | 8.86 | 0.25 [0.23 0.27] | 3.99 [3.81 4.16] | 0.78 [0.72 0.83] |
| | Emfit | 0.17 (3.89) [0.07 0.27] | -7.46 [-7.63 -7.29] | 7.8 [7.63 7.97] | 7.63 | 0.22 [0.19 0.24] | 2.64 [2.46 2.81] | 0.63 [0.52 0.75] |
| Wake | WSA | -0.56 (4.63) [-0.7 -0.43] | -9.64 [-9.88 -9.41] | 8.52 [8.29 8.75] | 9.08 | 0.31 [0.29 0.34] | 4.07 [3.83 4.3] | 0.64 [0.59 0.68] |
| | Emfit | 1.22 (7.67) [0.91 1.54] | -13.8 [-14.35 -13.26] | 16.25 [15.71 16.79] | 15.03 | 0.46 [0.42 0.5] | 5.82 [5.34 6.29] | 0.43 [0.33 0.53] |

Note. Metrics of agreement for Overall Heart rate of the devices (Withings sleep analyser [WSA], Emfit) and the estimates during sleep and wake against PSG electrocardiogram (ECG) estimates (beats per minute, bpm). The estimates are compared at 1 minute temporal resolution. The values shown are the mean followed by the (standard deviation) and [95% confidence interval]. The metrics include Bias – difference in measurement between the Device and PSG ECG (Device - PSG ECG); Minimum detectable change (MDC) – smallest detectable change independent of measurement error (half of Bland Altman agreement width); Standardized absolute difference (SAD) – directionless version of Cohen’s d; Mean absolute percentage error (MAPE) – mean error in measurement; Intraclass correlation with two-way random effects (ICC) – measures of measurement reliability.

**Supplementary Table 4. Breathing rate agreement measures in the laboratory (1 min)**

| Breathing rate agreement | | Bias [95% CI] | LoA Lower bound [95% CI] | LoA Upper bound [95% CI] | MDC | SAD [95% CI] | MAPE [95% CI] | ICC [95% CI] |
|---|---|---|---|---|---|---|---|---|
| Overall | WSA | -0.93 (2.32) [-0.96, -0.9] | -5.47 [-5.53, -5.42] | 3.61 [3.56, 3.67] | 4.54 | 0.64 [0.63, 0.66] | 9.86 [9.69, 10.03] | 0.36 [0.3, 0.41] |
| | Emfit | -0.18 (2.74) [-0.24, -0.12] | -5.54 [-5.65, -5.44] | 5.18 [5.08, 5.29] | 5.36 | 0.63 [0.61, 0.65] | 11.07 [10.74, 11.4] | 0.23 [0.17, 0.3] |
| | Somnofy | -0.47 (2) [-0.52, -0.43] | -4.38 [-4.46, -4.3] | 3.44 [3.36, 3.52] | 3.91 | 0.48 [0.46, 0.51] | 6.76 [6.54, 6.99] | 0.4 [0.32, 0.48] |
| Sleep | WSA | -1.01 (2.07) [-1.04, -0.98] | -5.08 [-5.13, -5.02] | 3.06 [3 3,.11] | 4.07 | 0.62 [0.6, 0.63] | 8.71 [8.56, 8.87] | 0.38 [0.32, 0.43] |
| | Emfit | -0.1 (2.52) [-0.16, -0.04] | -5.03 [-5.14, -4.92] | 4.83 [4.72, 4.94] | 4.93 | 0.6 [0.58, 0.63] | 9.6 [9.24, 9.96] | 0.25 [0.16, 0.34] |
| | Somnofy | -0.41 (1.73) [-0.45, -0.36] | -3.79 [-3.87, -3.71] | 2.98 [2.9, 3.06] | 3.39 | 0.44 [0.42, 0.47] | 5.88 [5.67, 6.1] | 0.44 [0.35, 0.52] |
| Wake | WSA | -0.69 (2.98) [-0.78, -0.6] | -6.53 [-6.68, -6.38] | 5.15 [5, 5.3] | 5.84 | 0.73 [0.7, 0.76] | 13.6 [13.11, 14.08] | 0.25 [0.2, 0.3] |
| | Emfit | -0.4 (3.25) [-0.54, -0.26] | -6.77 [-7, -6.53] | 5.97 [5.73, 6.21] | 6.37 | 0.71 [0.66, 0.75] | 15.04 [14.31, 15.77] | 0.19 [0.12, 0.25] |
| | Somnofy | -0.73 (2.78) [-0.88, -0.59] | -6.18 [-6.43, -5.93] | 4.71 [4.46, 4.97] | 5.45 | 0.63 [0.58, 0.68] | 10.15 [9.49, 10.8] | 0.29 [0.18, 0.4] |

Note. Metrics of agreement for Overall breathing rate and the estimates of the devices (Withings sleep analyser [WSA], Emfit and Somnofy) during sleep and wake against PSG respiratory inductance plethysmography (RIP) thorax estimates (cycles per minute, bpm). The values shown are the mean followed by the (standard deviation) and [95% confidence interval]. The metrics include Bias – difference in measurement between the Device and PSG RIP thorax estimate (Device – PSG RIP thorax); Lower and Upper bounds of the Bias; Minimum detectable change (MDC) – smallest detectable change independent of measurement error (half of Bland Altman agreement width); Standardized absolute difference (SAD) – directionless version of Cohen's d; Mean absolute percentage error (MAPE) – mean error in measurement; Intraclass correlation with two-way random effects (ICC) – measures of measurement reliability.

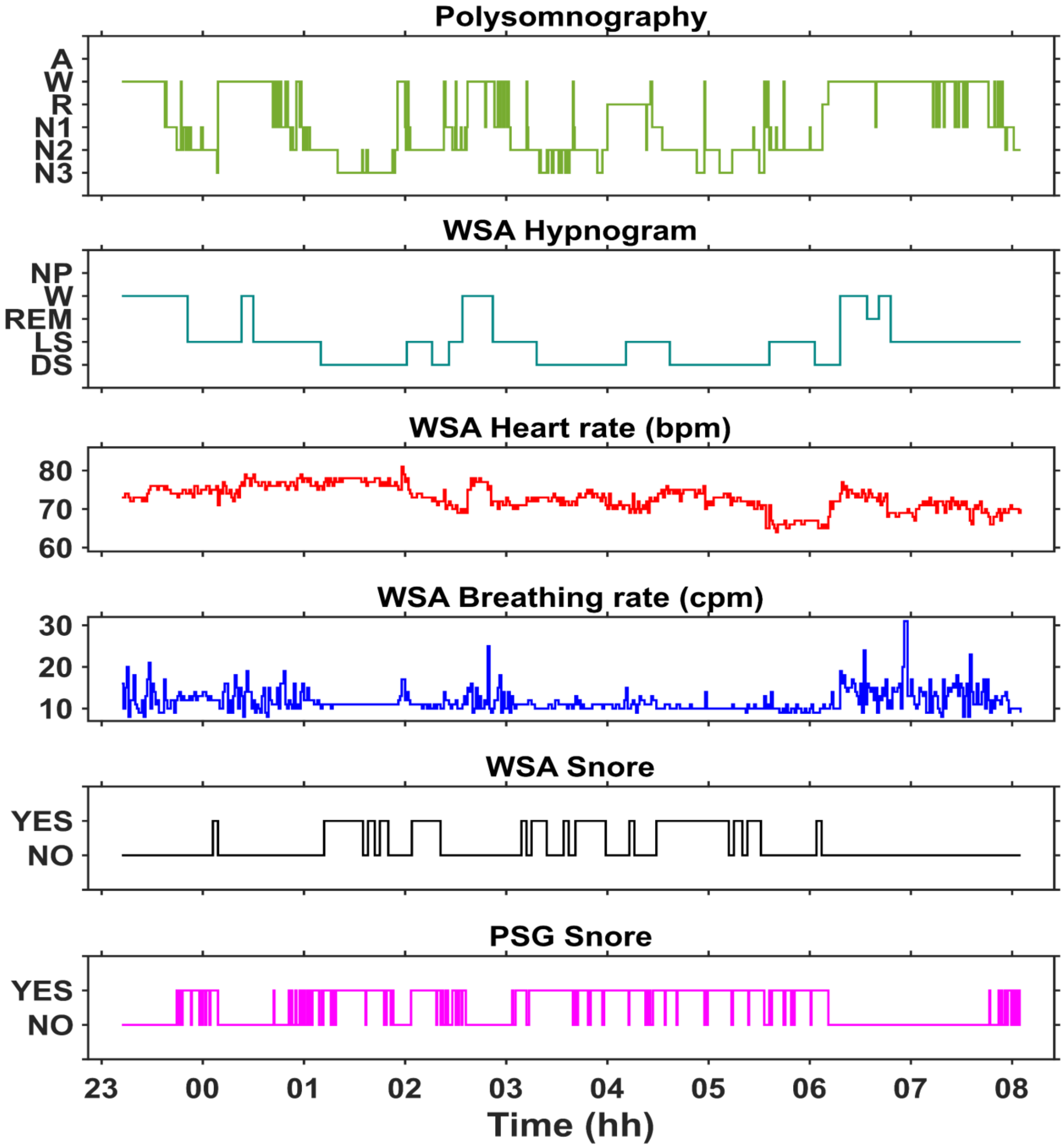


**Supplementary Figure 6. An example overnight time course of the data containing WSA snore and PSG snore.** The PSG consensus hypnogram, WSA hypnogram, WSA heart rate and breathing rate are also provided.

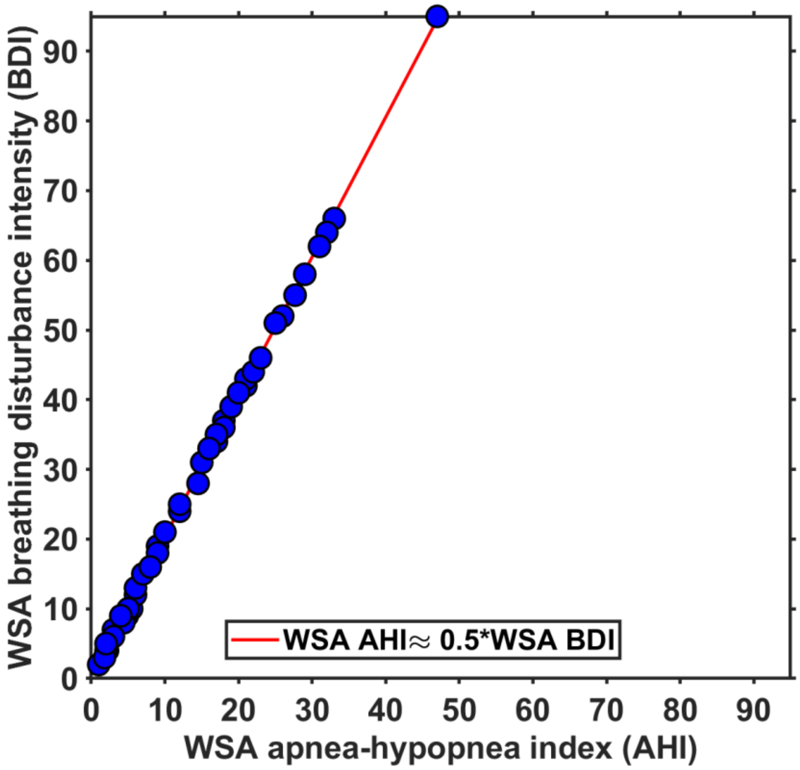


**Supplementary Figure 7. Relation between WSA apnea-hypopnea index (WSA AHI) and WSA breathing disturbance intensity (WSA BDI).** The linear fit is depicted by the red line.

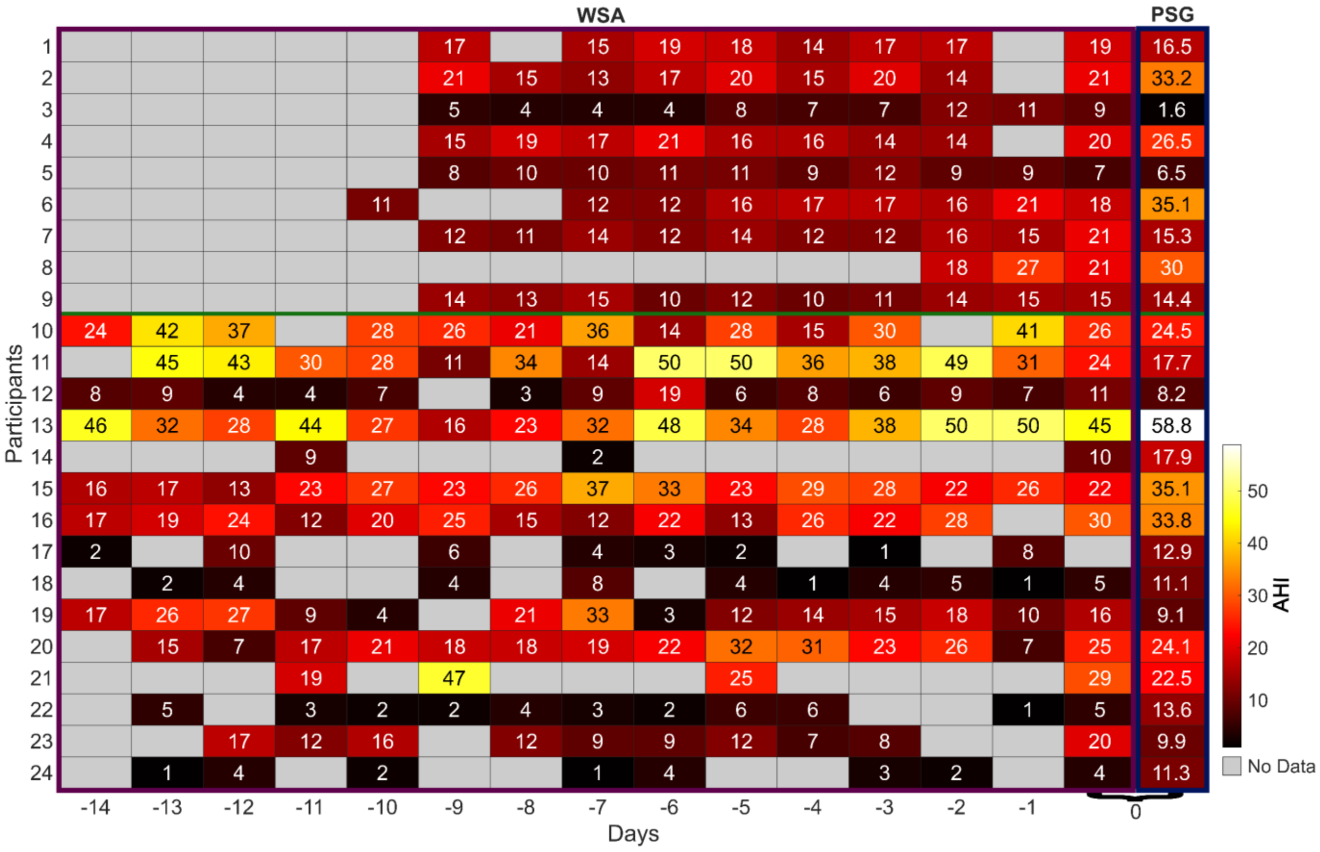


**Supplementary Figure 8. WSA AHI data at home.** The heatmap is plotted for the 24 participants for whom data were available both at home and in the laboratory and shows the night-to-night variability in WSA AHI. The WSA AHI is computed as half of the WSA BDI.
